# Quantifying longitudinal gait changes in ALS using wearable digital health technology metrics

**DOI:** 10.64898/2026.05.27.26354200

**Authors:** Katherine M. Burke, Narghes Calcagno, Sravan Mandepudi, Alan Premasiri, Kendall C. Hall, Fernando G. Vieira, James D. Berry, Marcin Straczkiewicz

## Abstract

Wearable digital health technologies may complement traditional gait assessments in amyotrophic lateral sclerosis (ALS) by sensitively capturing real-world mobility changes. In this study, we validated six digital gait metrics derived from ankle-worn sensors in a natural history cohort of 182 individuals with ALS. Investigated metrics correspond to various aspects of gait, including volume, speed, intensity, similarity, variability, and fragmentation. Longitudinal analyses showed significant declines in step count, peak cadence, stride intensity, and stride similarity, with increasing stride duration variability and walking fragmentation over 52 weeks. Many participants exhibited greater relative change in the gait metrics than the self-reported ALS Functional Rating Scale-Revised (ALSFRS-RSE). Stratified analyses revealed that digital metrics captured significant functional decline even in participants with stable walking scores on the ALSFRS-RSE. These findings support the potential utility of these metrics for disease monitoring in ALS clinical care and trials.

## 1. Introduction

People living with amyotrophic lateral sclerosis (plwALS) often experience progressive gait dysfunction, that reduces mobility, increases fall risk, and may adversely affect social connectedness and quality of life.^1,2^ Falls are common in this population, and head trauma resulting from falls accounts for approximately 1.7% of deaths among ambulatory plwALS.^3^ Given both the prevalence and the clinical consequences of gait impairment, there is a critical need for effective tools to monitor gait changes in this population.

Gait abnormalities in plwALS are complex and can be characterized by variables such as increased stride variability, decreased single-limb stance time, reduced step length, and lower cadence.^4,5^ These characteristics are quantifiable, change over time, and reflect both disease progression and clinically important functional status, yet traditional clinical assessments rely only on subjective observations and infrequent in-person evaluations.^6,7^ The ALS Functional Rating Scale-Revised (ALSFRS-R), one of the most commonly used endpoints in ALS clinical trials, captures walking ability using a single ordinal item, which may limit its sensitivity to subtle yet clinically meaningful changes over time.^8^ In clinical practice, gait can be directly assessed using timed tasks, such as the 10-Meter Walk, the Timed Up and Go, or the 6-Minute Walk Tests.^9–11^ During these assessments, clinicians must simultaneously ensure patient safety, time the task, and observe gait quality, limiting the objectivity, practicality, and granularity of these evaluations.

Wearable digital health technologies (wDHTs), including wrist and ankle-worn devices with inertial measurement units (IMUs), enable frequent and objective measurement of gait in naturalistic settings, and offer the potential to generate digital gait endpoints that are being shown to be more responsive to longitudinal change than traditional outcome measures.^12^ Prior work has demonstrated the feasibility of quantifying gait using wDHT-derived metrics such as step counts and cadence in ALS and related neurodegenerative diseases.^13,14^ Many promising metrics remain insufficiently validated for use as clinical or trial endpoints, particularly with respect to longitudinal performance and clinical relevance.^5^

The aim of this study was to establish clinical validation for digital gait metrics derived from a wDHT worn at the ankle to capture both the quantity and quality of gait in plwALS. In this work, we utilized a previously validated step-counting algorithm for plwALS,^13^ and derived additional gait metrics from the accelerometer data, including peak cadence, step intensity, stride pattern similarity, stride duration variability, and walking fragmentation. By quantifying changes in the volume, speed, intensity, variability, consistency, and fragmentation of gait during continuous real-world monitoring, this work will provide a more comprehensive assessment of gait and prepare us to incorporate digital gait metrics as outcome measures in ALS trials.

## 2. Methods

### 2.1. Participants and data sources

Data were collected by the ALS Therapy Development Institute (ALS TDI) as part of the ALS Research Collaborative (ARC).^15^ To date, the study has enrolled over 1500 people living with ALS and assembled a rich dataset including self-reported ALSFRS-R (ALSFRS-RSE) scores together with digital physiologic data, skin biopsies, genome sequences, plasma samples, and speech recordings, with a subset contributing accelerometer measurements.^6^

For this analysis, we used accelerometer data from wDHTs and ALSFRS-RSE scores collected between 2014 and 2023 in the ALS TDI ARC study.^6^ The accelerometer data were collected using an ActiGraph GT3X+ sensor (Ametris, Pensacola, FL) worn at the left ankle typically for 7 to 11 days every three to four weeks. Participants were encouraged to wear the device during short prescribed exercises completed three times over the course of a recording period, and for as long as they wanted for the remainder of recording period.^6^ This liberal data collection regime was implemented to improve participant retention in this natural history study. The devices collected continuous triaxial accelerometer measurements with a sampling frequency of 30 Hz and a dynamic range of ± 6 g (gravitational units).^6,16^

The ALSFRS-RSE is a patient-reported 12-item survey with each item scored 0—4 points, with higher scores indicating better functioning. Survey scores were collected every four to six weeks using a web-based platform, as described previously.^6,16^

### 2.2. Digital gait metrics

Gait metrics were extracted from raw accelerometer data collected by the ankle-worn devices using a validated statistical method developed by our team.^13^ In short, the method leverages foot contact events occurring during the initial phase of gait, which are reflected in the accelerometer signal as high-amplitude spikes. To extract the timing of these events, we applied a continuous wavelet transform with a generalized Morse wavelet as a mother wavelet and a narrow frequency response. Given the repetitive nature of walking, we identified gait strides as two consecutive foot contacts occurring at least 0.85 and no more than 2.5 seconds apart. Step counts were calculated as twice the number of detected strides. To restrict the analysis to periods of physiologically plausible walking only, we further considered walking bouts consisting of at least three closely spaced foot contacts, corresponding to at least two consecutive strides.

In this analysis, we investigated six gait-related digital metrics derived at the daily level and averaged across each monitoring period: 1) step counts, 2) peak cadence, 3) stride intensity, 4) stride duration variability, 5) stride pattern similarity, and 6) walking fragmentation (**Table 1**). Briefly, step counts are the total number of daily steps. Peak cadence is the greatest steps per minute over two consecutive minutes in a day, averaged across the monitoring period. Stride intensity is the stride-specific vertical amplitude of acceleration, i.e., axis parallel to participant’s shin. Stride pattern similarity was calculated as a piecewise correlation between time-normalized consecutive stride patterns derived from acceleration collected in the vertical-axis, averaged for all consecutive stride pattern comparisons. Stride duration variability was calculated as the standard deviation of stride-to-stride duration differences. Finally, walking fragmentation describes the ratio of the number of walking bouts to total walking minutes each day. It is calculated as the reciprocal of the mean duration of each walking bout, consistent with previous work evaluating active-to-sedentary behavior transitions.^17^ These metrics were selected to capture complementary dimensions of gait and to provide insights into both gait quantity and quality across stages of ALS progression. Their units, operational definitions, and representative gait domains are provided in **Table 1**; their graphical interpretation is presented in **Figure 1**.

**Table 1.** Digital gait metrics derived from accelerometer data and investigated in this study. A valid monitoring period consisted of at least 3 days, each with at least 16 hours of sensor wear-time. Each metric is derived at the daily level and then averaged across the monitoring period.

| Digital gait metric | Units | Description | Gait aspect |
| --- | --- | --- | --- |
| Step counts | Steps | The sum of twice the number of detected strides | Volume |
| Peak cadence | Steps · min<sup>-1</sup> | The greatest number of steps observed across 2 consecutive minutes | Speed |
| Stride intensity | g (gravitational units) | Average stride acceleration amplitude | Intensity |
| Stride pattern similarity | Unitless | Average of each of the piecewise correlations between time-normalized consecutive stride patterns | Similarity |
| Stride duration variability | Seconds | Standard deviation of stride-to-stride duration differences | Variability |
| Walking fragmentation | bouts $\cdot \text{min}^{-1}$ | Ratio of the number of walking bouts to the total number of walking minutes | Fragmentation |

**Figure 1.**
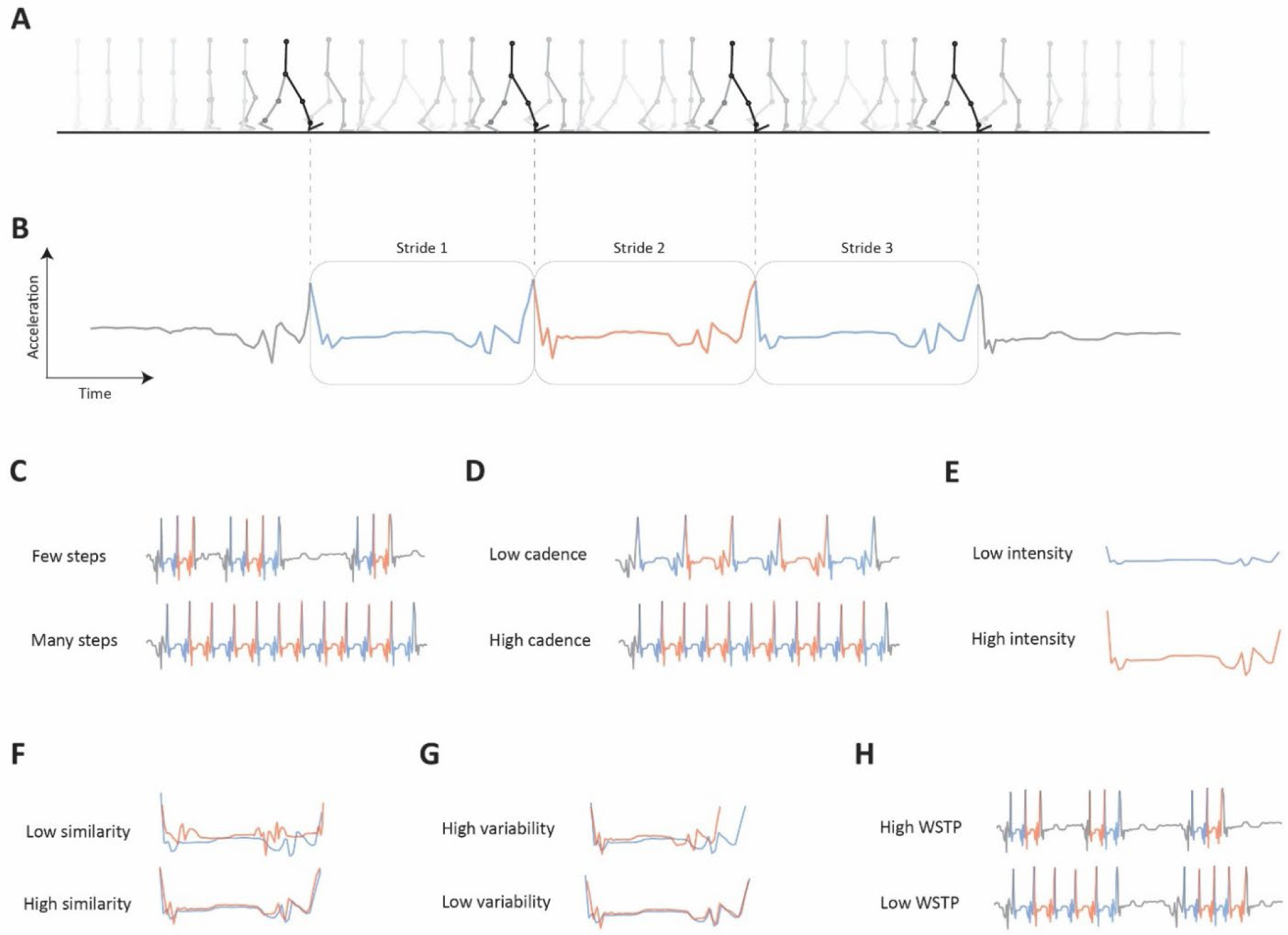
Schematic representation of gait cycles, corresponding vertical-axis accelerometer signal, and derived digital gait metrics. Repetitive cycles (**A**) result in local spikes in acceleration related to left-foot ground contact and indicate the beginning and end of a detected stride (two steps) (**B**). We derived six metrics: step counts (**C**), peak cadence (**D**), stride intensity (**E**), stride pattern similarity (**F**), stride duration variability (**G**), and walking fragmentation (**H**). To ease interpretation, panels **C-H** show examples of lower and higher values of each metric. In panels **B-H**, blue and red colors were used to differentiate signal fragments corresponding to subsequent strides, while black color was used to indicate periods without detected strides.

### 2.3. Statistical analysis

ALSFRS-RSE scores were included in the analysis if they were collected within 7 days before or after the corresponding accelerometer data collection period. A valid monitoring period was defined as one containing at least three valid wear days. Days were considered valid if they included at least 16 hours of sensor wear-time, as determined using the algorithm by Choi et al^18^ and consistent with previously validated wear-time cutoffs.^19–21^ Digital gait metrics were calculated at the daily level and then averaged across all valid days within each data collection bout.

The analytic sample comprised digital gait metrics and ALSFRS-RSE scores from participants who completed at least two surveys and contributed at least two valid accelerometer data collection monitoring periods within 52 weeks of enrollment. This time window was imposed to reduce the influence of slow progressors on model estimates and to approximate the design of a hypothetical clinical trial.^22^ Importantly, our analysis was restricted to plwALS who were ambulatory at enrollment, as defined by their first response to question 8 (Q8; “ability to walk”). In this work, we focused on participants reporting their score as either 4, 3, or 2, which corresponded to normal walking, early ambulation difficulties, or walking with assistance, respectively.

To summarize the dataset, we computed the total follow-up time, weeks with accelerometer data, and time between collection bouts. For survey data, we computed the total number and the average time between survey completions. Statistics were computed for each participant and reported on a population level using mean and standard deviation (SD).

Depending on the aspect of investigation, our analysis leveraged different survey scores: ALSFRS-RSE total score (Q1—12; score 0-48) was used to examine baseline correlation with digital gait metrics (**Section 3.2.1**), association with digital gait metrics (**Section 3.2.2**), and association with time elapsed since baseline (**Sections 3.3.1** and **3.3.2**); gross motor subdomain score (Q7—9; score 0-12) was used to examine baseline correlation with digital gait metrics and association with time elapsed since baseline; walking ability score (Q8; score 0-4) was used to stratify participants by self-reported disease status (**Section 3.3.3**) and to estimate minimal detectable change (MDC) (**Section 3.4**). Finally, bulbar (Q1—3; score 0-12), fine motor (Q4—6; score 0-12), and respiratory (Q10—12; score 0-12) subdomain scores were used to examine baseline correlations with digital gait metrics (**Section 3.2.1**).

### Associations between digital gait metrics and ALSFRS-RSE scores

Baseline correlations were evaluated with Pearson correlation using the first valid pair of digital metrics and survey scores. Correlation coefficients were interpreted as follows^23^:

- <0 – 0.20: very weak correlation
- 0.20–0.40: weak correlation
- 0.40–0.60: moderate correlation
- 0.60–0.80: strong correlation
- >0.80: very strong correlation

Given repeated observations within individuals, the associations between digital metrics and survey responses were further examined using linear mixed-effects models (LMMs). Each of the models specified survey response (either total (Q1—12) or gross motor (Q7—9)) as an outcome and the digital metric (one of six) as a fixed effect. To account for participant-level baseline values and disease progression rate, they also included random intercept and random slope. These models evaluated whether and how digital gait metrics correlate with changes in survey responses.

### Longitudinal changes in digital gait metrics

To evaluate longitudinal changes in the digital gait metrics, separate LMMs were fitted with the digital metric (one of six) as an outcome, time since baseline as a fixed effect, and participant-specific random slope (at the weekly level) and random intercept. Similar LMMs were also fitted for total (Q1—12) or gross motor (Q7—9) scores as outcomes. Population-level relative change was calculated as a ratio between population-level slope and intercept, and expressed as the percent change over 52 weeks. To compare trajectories across outcomes measured on different scales, we also divided the fixed effect of time by the overall standard deviation of each outcome, and provided this standardized rate of change in units of SD per 52 weeks.

Participant-level intercepts and slopes were derived by combining the fixed-effect estimates from the LMMs with the corresponding participant-level random effects. The participant-specific relative change was then calculated as the ratio of the participant-specific slope to the participant-specific intercept. Both population- and participant-level relative changes were multiplied by 100 yielding an estimate of percent change in the outcome per unit increase in time, i.e., weekly, and multiplied by 52 weeks to estimate the percent change over 52 weeks.

### Stratification by walking ability and decline in ambulation

To understand how digital metrics change over time in relation to initial gait status and disease progression, we stratified participants into groups based on their initial gait status (scores of 4, 3, or 2 on Q8 of the ALSFRS-RSE, since participants were not enrolled if they had a score of 0 or 1), and by whether their walking score remained stable or improved (indicating no decline, non-progressors) versus a decrease by one or more points on Q8 of the ALSFRS-RSE over the 52 weeks (indicating a decline, progressors). Separate LMMs for each of the stratified groups were fitted using data from participants within each of these groups, with each digital metric as the outcome, time elapsed from baseline as a fixed effect, and participant-specific random slopes and intercepts. This analysis allowed us to estimate rate of change (slope) separately in each metric for individuals with and without gait deterioration captured by the ALSFRS-RSE. Differences in longitudinal change between groups were assessed by including a group interaction term. The statistical significance of this interaction term was used to determine whether the slopes differed between groups.

### Reliability and minimal detectable change

Minimal detectable change (MDC) was calculated to determine the smallest change exceeding measurement error at 95% confidence using the standard error of measurement (SEM), derived from the standard deviation of the metric and its test–retest reliability estimated using intraclass correlation coefficient (ICC, two-way random effects, absolute agreement).^24,25^ Data were drawn from two consecutive recording periods in which participants had both a valid week of accelerometer data and reported no change in walking ability on Q8 of the ALSFRS-RSE. Participants could contribute more than one pair of observations, provided they met these criteria. MDC was estimated for individuals with different walking abilities, stratified by ambulatory status on Q8 of the ALSFRS-RSE, with scores of 4, 3, and 2 corresponding to normal walking, experiencing early ambulation difficulties, or walking with assistance, respectively. ICC values were interpreted as the following^25^:

- <0.50: poor reliability
- 0.50 to <0.75: moderate reliability
- 0.75 to <0.9: good reliability
- ≥0.90: excellent reliability

We accounted for multiple tests using the Bonferroni for the six digital gait metrics; based on a nominal type I error rate of 0.05 and with six tests, the resulting adjusted cutoff *p*-value for statistical significance was estimated as equal to 0.008.^26^ Model assumptions were evaluated through visual inspection of residual histograms, probability plots, and residual-versus-fitted plots.

To align with common clinical trial follow-up periods, data were included from a predefined time window of the initial 52 weeks since enrollment. We also considered shorter time windows corresponding to initial 26 and 13 weeks. Results for the shorter follow-up periods are provided in **Supplementary Materials**.

## 3. Results

### 3.1. Demographics, compliance, functional status

Out of 349 participants with any accelerometer data collected, 12 were non-ambulatory (reported 1 or 0 on ALSFRS-R Q8) at baseline, while additional 155 did not collect data with sufficient wear-time during two or more monitoring bouts. Therefore, our analytic sample included data from 182 participants. Participants were predominantly white (97%) males (65%) with a mean age of 57 years and baseline total ALSFRS-RSE score of 40.41 (**Table 2**).

**Table 2.**
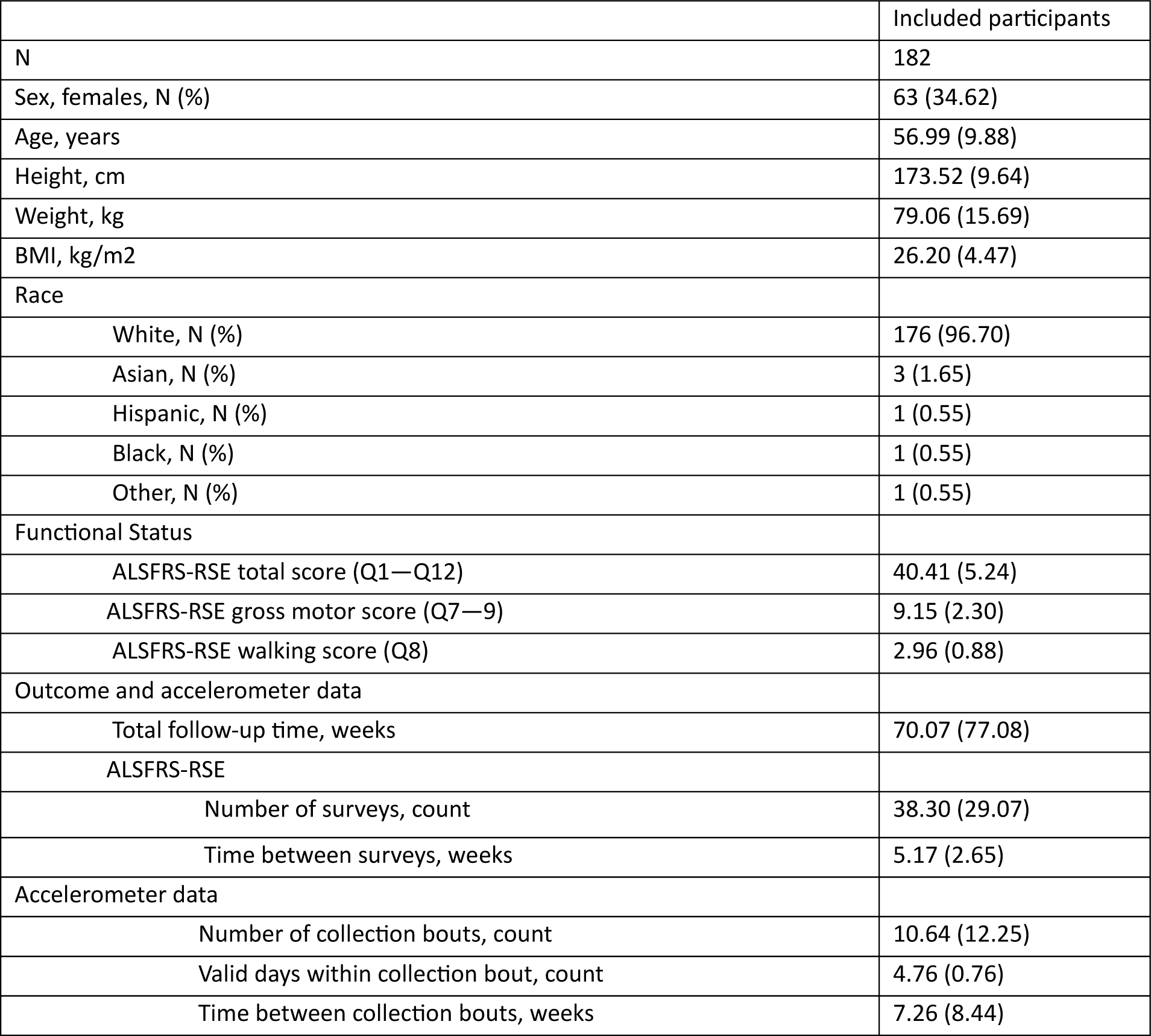
Baseline demographics and descriptive statistics for ALSFRS-RSE and accelerometer data in the analytic sample, presented as mean (SD) unless otherwise noted.

|  | Included participants |
| --- | --- |
| N | 182 |
| Sex, females, N (%) | 63 (34.62) |
| Age, years | 56.99 (9.88) |
| Height, cm | 173.52 (9.64) |
| Weight, kg | 79.06 (15.69) |
| BMI, kg/m<sup>2</sup> | 26.20 (4.47) |
| Race |  |
| White, N (%) | 176 (96.70) |
| Asian, N (%) | 3 (1.65) |
| Hispanic, N (%) | 1 (0.55) |
| Black, N (%) | 1 (0.55) |
| Other, N (%) | 1 (0.55) |
| Functional Status |  |
| ALSFRS-RSE total score (Q1—Q12) | 40.41 (5.24) |
| ALSFRS-RSE gross motor score (Q7—9) | 9.15 (2.30) |
| ALSFRS-RSE walking score (Q8) | 2.96 (0.88) |
| Outcome and accelerometer data |  |
| Total follow-up time, weeks | 70.07 (77.08) |
| ALSFRS-RSE |  |
| Number of surveys, count | 38.30 (29.07) |
| Time between surveys, weeks | 5.17 (2.65) |
| Accelerometer data |  |
| Number of collection bouts, count | 10.64 (12.25) |
| Valid days within collection bout, count | 4.76 (0.76) |
| Time between collection bouts, weeks | 7.26 (8.44) |

### 3.2. Associations with ALSFRS-RSE scores

#### 3.2.1 Cross-sectional correlations

Baseline associations between each of the digital gait metrics and the ALSFRS-RSE (total and each subdomain) revealed significant positive correlations with the total score and gross motor subdomain for every gait metric except for stride duration variability and walking fragmentation, which demonstrated significant negative associations (**Figure 2**). Specifically, higher step counts, faster peak cadence, greater intensity, and greater stride pattern similarity were associated with higher ALSFRS-RSE total and gross motor subdomain scores. Conversely, higher stride duration variability and higher walking fragmentation were associated with lower gross motor and total ALSFRS-RSE scores.

**Figure 2.**
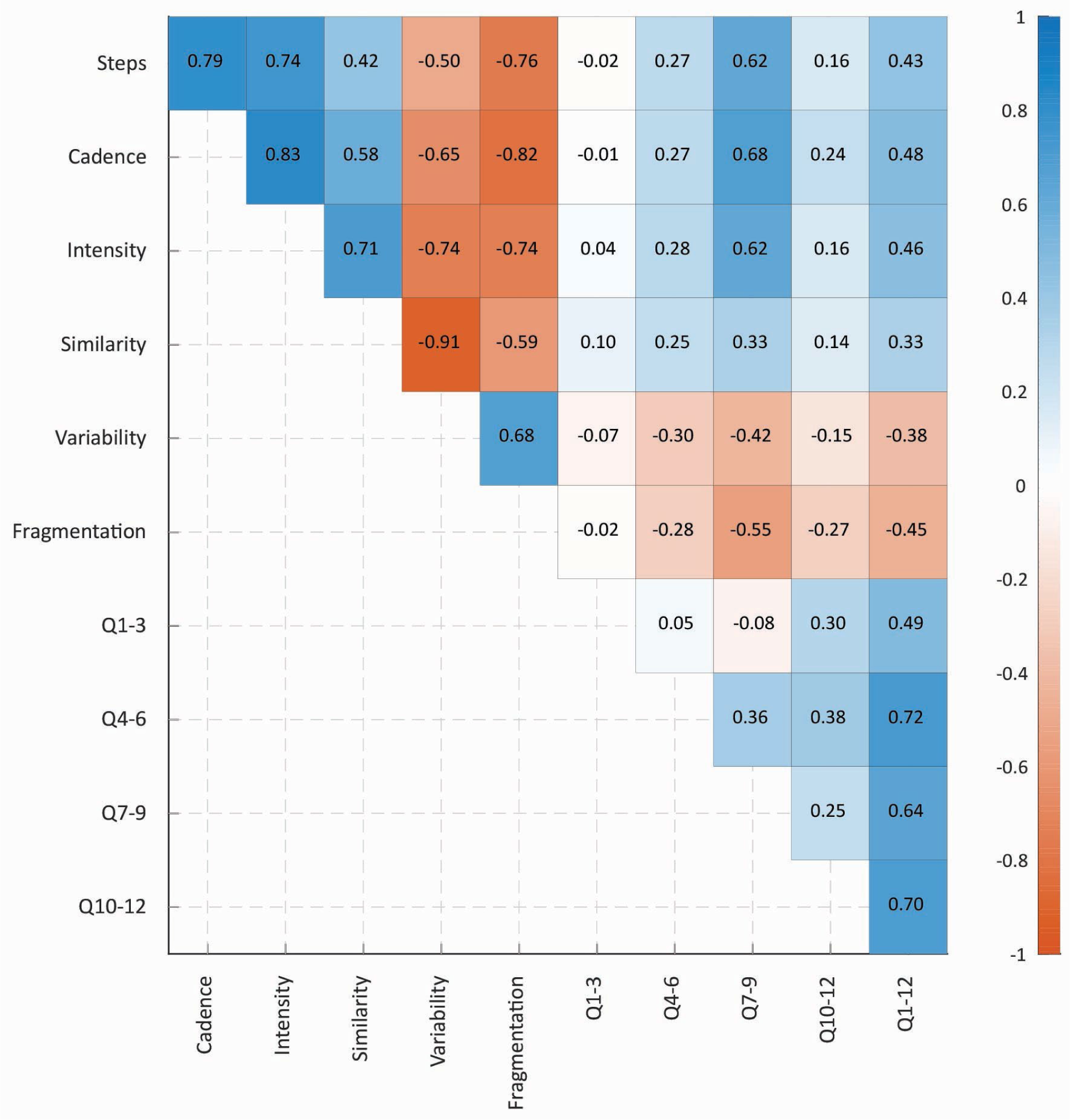
Correlation matrix between investigated digital gait metrics and ALSFRS-RSE survey responses observed at baseline. Pearson correlation coefficients were calculated for six gait metrics and four survey subdomains, bulbar (Q1–3), fine motor (Q4–6), gross motor (Q7–9), and respiratory (Q10–12), as well as total score (Q1-12).

Correlations with gross motor subdomain of the ALSFRS-RSE were moderate to strong for peak cadence (Rho = 0.68), followed by step counts (0.62), stride intensity (0.62) walking fragmentation (-0.55), and stride duration variability (-0.42), and weak for stride pattern similarity (0.33). Very weak to weak correlations were observed between the digital metrics and bulbar (Q1—3), respiratory (Q10—12) and fine motor function (Q4—6). These findings support the convergent and divergent validity of each of the gait metrics with the respective sub-domains of the ALSFRS-RSE.

Notably, strong to very strong correlation coefficients (Rho <-0.6 or >0.6) were observed between several digital gait metrics, with the highest noted between stride pattern similarity and stride duration variability (-0.91), between peak cadence and stride intensity (0.83), and between peak cadence walk-to-sedentary transition probability (-0.82). This suggests that digital metrics measure independent but highly correlated features of gait.

#### 3.2.2 Longitudinal mixed-effects associations

Separate LMMs were fit to examine the relationship between digital gait metrics and survey responses. Positive associations were observed between the total ALSFRS-RSE scores and four metrics: step counts (mean 148.7 [119.6, 178.0] steps), peak cadence (2.81 [2.35, 3.27] steps ·min^-1^), stride intensity (0.031 [0.025, 0.036] g), and stride pattern similarity (0.012 [0.008, 0.016]) for a one point increase in total ALSFRS-R scores. While negative associations were noted for stride duration variability (-0.010 [-0.013, -0.007] seconds) and fragmentation (-0.016 [-0.020, -0.012] bouts ·min^-1^). All associations were statistically significant (*p*<0.001). These findings indicate that participants with higher overall function, as measured by the ALSFRS-RSE, walked more and with faster cadence, while their strides were more intense and similar. Our results also suggest that stride duration of higher functioning participants was less variable and walking was less fragmented (**Figure 3**).

**Figure 3.**
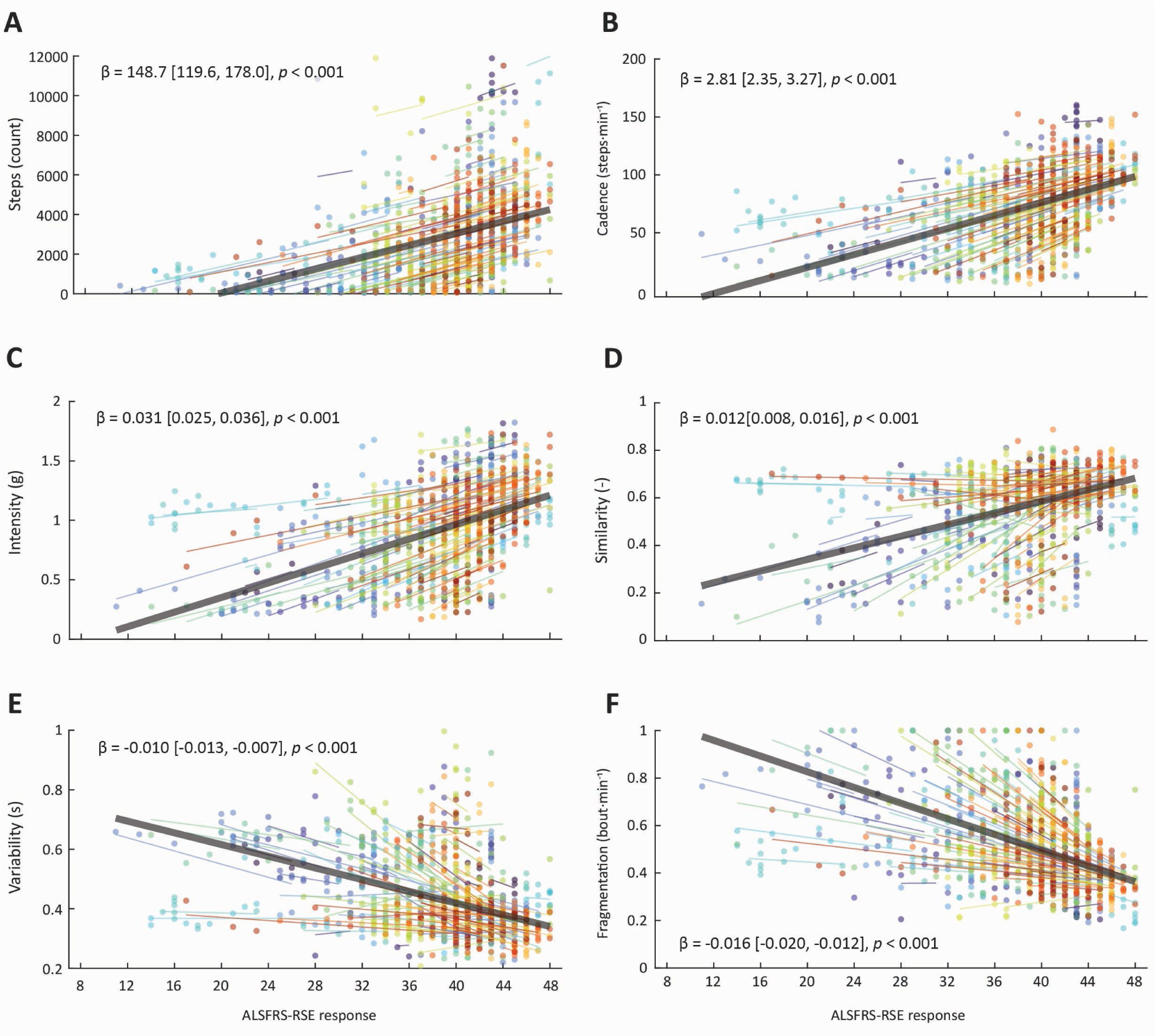
The association between digital gait metrics and ALSFRS-RSE total score (Q1—12) estimated using LMMs. Each panel represents one metric: step counts (**A**), peak cadence (**B**), stride intensity (**C**), stride pattern similarity (**D**), stride duration variability (**E**), and walking fragmentation (**F**). In each plot, the colored dots represent observed values, colored lines represent participants’ conditional means, while black lines represent population mean. Points and lines are color-coded by participant (the colors are fixed across the panels).

### 3.3. Longitudinal changes in digital gait metrics

#### 3.3.1 Population-level trajectories

Separate LMMs were fit to examine the association between digital gait metrics, survey responses, and time elapsed since baseline. The models revealed a significant decline in step counts, peak cadence, stride intensity, and stride pattern similarity over time, and an increase in stride duration variability and walking over time (all *p*<0.001). This indicates that over time participants made fewer steps with a slower cadence, lower stride intensity, and decreased stride similarity, while their stride duration was more variable and walking bouts were more fragmented (**Table 3**). Expectedly, total and gross motor sub-domain scores on the ALSFRS-RSE demonstrated significant decline over time.

**Table 3.** Model coefficients for digital gait metrics and functional scores, represented by the total and gross motor sub-domain scores on the ALSFRS-RSE. Model slopes express weekly change in digital metrics and survey scores. Standardized and relative changes are expressed in SD and percentage change per 52 weeks, respectively.

| Outcome | Intercept | Slope | R-squared | Standardized change (SD) | Relative change (%) |
| --- | --- | --- | --- | --- | --- |
| Steps | 3280 [2953, 3608] | -22.88 [-29.03, -16.73] | 0.849 [0.854, 0.884] | -0.506 | -36.3 |
| Cadence | 79.92 [75.95, 83.88] | -0.43 [-0.53, -0.36] | 0.792 [0.798, 0.864] | -0.735 | -27.8 |
| Intensity | 1.01 [0.962, 1.06] | -0.005 [-0.006, -0.003] | 0.844 [0.862, 0.894] | -0.678 | -23.8 |
| Similarity | 0.599 [0.579, 0.619] | -0.002 [-0.002, -0.001] | 0.725 [0.769, 0.837] | -0.584 | -15.1 |
| Variability | 0.405 [0.389, 0.4219] | 0.001 [0.001, 0.002] | 0.825 [0.838, 0.874] | 0.570 | 17.5 |
| Fragmentation | 0.464 [0.444, 0.484] | 0.003 [0.002, 0.003] | 0.700 [0.721, 0.771] | 0.741 | 28.5 |
| ALSFRS-RSE gross motor score (Q7—9) | 9.18 [8.84, 9.52] | -0.039 [-0.047, -0.031] | 0.969 [0.971, 0.973] | -0.833 | -22.2 |
| ALSFRS-RSE total score (Q1—12) | 40.44 [39.66, 41.22] | -0.110 [-0.130, -0.091] | 0.975 [0.977, 0.977] | -0.941 | -14.2 |

Note: For each model estimated $p < 0.001$ .

At the population-level, four out of six of the digital metrics (step counts, peak cadence, stride intensity, and fragmentation) experienced greater percent relative change compared to gross motor subdomain score, and the same was the case for all metrics when compared to ALSFRS-RSE total score. Although the standardized change was greatest for the ALSFRS-RSE total score (-0.941 SD) over the 52 weeks.

#### 3.3.2 Participant-level trajectories

To characterize participant-level differences in the relative rates of change across gait metrics and ALSFRS-RSE scores, individual point estimates of weekly relative change were calculated for each participant. On the participant-level, of the 182 participants, 164 (90%) exhibited a greater percent relative change in step counts, 149 (82%) in peak cadence, 140 (77%) in stride intensity, 133 (73%) in fragmentation, 102 (56%) in stride duration variability, and 86 (47%) in stride similarity, compared to the total ALSFRS-RSE scores over 52 weeks (**Figure 4**).

**Figure 4.**
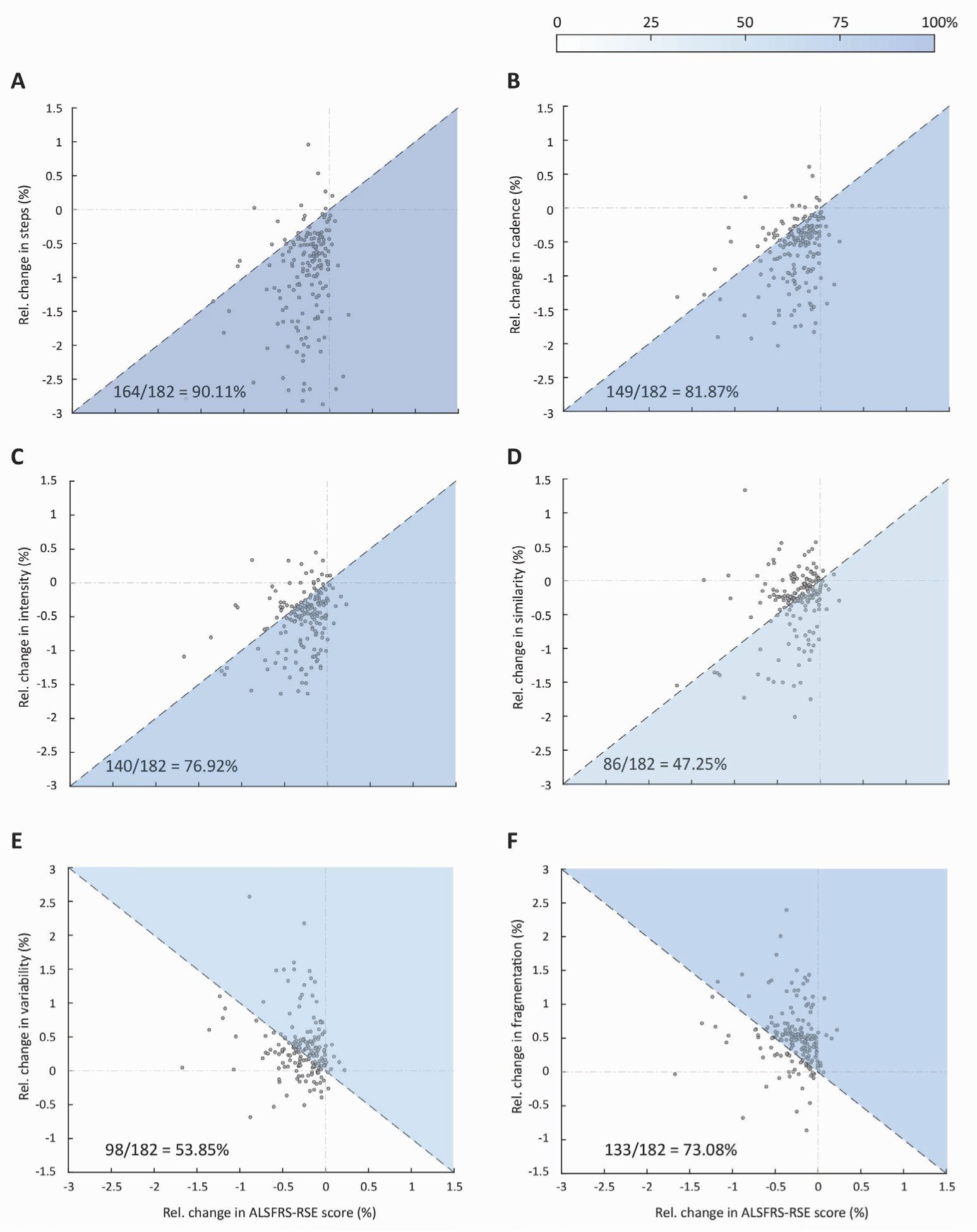
Point estimates for participant-specific relative change per week in digital gait metrics compared to corresponding relative change in ALSFRS-RSE total score within a 52-week window. Each panel corresponds to one metric: step counts (**A**), peak cadence (**B**), stride intensity (**C**), stride pattern similarity (**D**), stride duration variability (**E**), and walking fragmentation (**F**). Each point represents the relative change in the digital metric and ALSFRS-RSE total score in one participant; dashed lines correspond to identical relative change observed in the digital gait metric and the ALSFRS-RSE total score. The shaded sections highlight individuals with relative change in the digital metric exceeding the relative change in their ALSFRS-RSE total score. Tones of blue indicate the percentage of participants with greater relative change exhibited by a digital metric, with darker blues indicating higher percentage.

#### 3.3.3 Stratification by self-reported ambulatory status and its change

Separate LMMs were fitted to examine the longitudinal trajectory of digital metrics in each ambulatory status group at baseline (Q8 scores of 4, 3, or 2), stratified by non-progressors and progressors (**Table 4**). Non-progressors with normal gait at baseline (Q8 = 4) did not demonstrate significant change in any of the digital metrics using the adjusted *p* value of 0.008. In contrast, progressors who reported normal gait at baseline exhibited a significant decrease in peak cadence (*p*=0.001), stride intensity (*p*=0.003), and steps counts (*p*=0.005). The rate of change over the 52 weeks, however, was not statistically different between these groups for any of the digital gait metrics (**Figure 5**).

**Table 4.** Comparison between longitudinal changes in gait metrics between individuals reporting no decline (non-progressors) and those reporting decline by at least one point (progressors) on ALSFRS-RSE Q8. Model slopes express weekly change in digital metrics and survey scores. Standardized and relative changes are expressed in SD and percentage change per 52 weeks, respectively.

| Digital metric | Intercept | Slope | p-value | R-squared | Standardized change (SD) | Relative change (%) | Intercept | Slope | p-value | R-squared | Standardized change (SD) | Relative change (%) | Slope difference p-value |
| --- | --- | --- | --- | --- | --- | --- | --- | --- | --- | --- | --- | --- | --- |
|  | **Non-progressors, baseline Q8 = 4 (n=32)** |  |  |  |  |  | **Progressors, baseline Q8 = 4 (n=32)** |  |  |  |  |  |  |
| Steps | 4805<br>[4128, 5482] | -13.80 [-25.14, -2.45] | 0.017 | 0.563 | -0.333 | -14.924 | 4800<br>[3924, 5675] | -21.60 [-36.60, -6.60] | **0.005** | 0.743 | -0.415 | -23.4 | 0.454 |
| Cadence | 95.85<br>[89.72, 102] | -0.170 [-0.303, -0.037] | 0.012 | 0.429 | -0.504 | -9.204 | 98.59<br>[92.66, 104.5] | -0.371 [-0.586, -0.156] | **0.001** | 0.513 | -0.777 | -19.604 | 0.283 |
| Intensity | 1.19 [1.12, 1.27] | -0.001 [-0.001, 0.000] | 0.081 | 0.496 | -0.352 | -2.184 | 1.219<br>[1.155, 1.283] | -0.003 [-0.005, -0.001] | **0.003** | 0.643 | -0.700 | -13.364 | 0.057 |
| Similarity | 0.614<br>[0.587, 0.641] | -0.000 [-0.000, 0.000] | 0.680 | 0.589 | -0.098 | -0.364 | 0.623<br>[0.595, 0.651] | -0.000 [-0.001, 0.000] | 0.851 | 0.341 | -0.032 | -0.676 | 0.726 |
| Variability | 0.376<br>[0.358, 0.395] | 0.000 [-0.000, 0.000] | 0.267 | 0.549 | 0.044 | 0.988 | 0.374<br>[0.354, 0.395] | -0.000 [-0.001, 0.000] | 0.690 | 0.197 | 0.058 | 1.248 | 0.587 |
| Fragmentation | 0.396<br>[0.368, 0.424] | 0.001 [-0.000, 0.001] | 0.114 | 0.353 | 0.350 | 6.812 | 0.397<br>[0.367, 0.426] | 0.001 [0.000, 0.002] | 0.025 | 0.383 | 0.468 | 13.312 | 0.424 |
|  | **Non-progressors, baseline Q8 = 3 (n=23)** |  |  |  |  |  | **Progressors, baseline Q8 = 3 (n=27)** |  |  |  |  |  |  |
| Steps | 4560<br>[3781,<br>5339] | -16.25 [-<br>25.45,-<br>7.06] | **0.001** | 0.554 | -0.402 | -18.512 | 3134<br>[2615,<br>3653] | -36.23 [-<br>47.30,-<br>25.15] | **<0.001** | 0.839 | -1.164 | -60.112 | **<0.001** |
| Cadence | 97.9 [91.4,<br>104.4] | -0.339 [-<br>0.531,-<br>0.147] | **0.001** | 0.611 | -0.986 | -17.992 | 83.59<br>[76.46,<br>90.71] | -0.530 [-<br>0.727,-<br>0.332] | **<0.001** | 0.736 | -1.144 | -32.968 | 0.394 |
| Intensity | 1.22 [1.12,<br>1.32] | -0.004 [-<br>0.010,<br>0.001] | 0.110 | 0.636 | -0.848 | -18.876 | 1.09<br>[0.992,<br>1.18] | -0.006 [-<br>0.009,-<br>0.004] | **<0.001** | 0.583 | -1.099 | -30.628 | 0.402 |
| Similarity | 0.662<br>[0.632,<br>0.692] | -0.001 [-<br>0.003,<br>0.000] | 0.112 | 0.933 | -0.716 | -9.932 | 0.651<br>[0.625,<br>0.676] | -0.002 [-<br>0.004,-<br>0.001] | **0.004** | 0.660 | -1.145 | -19.864 | 0.348 |
| Variability | 0.348<br>[0.329,<br>0.366] | 0.001 [-<br>0.000,<br>0.002] | 0.221 | 0.512 | 0.693 | 11.128 | 0.358<br>[0.334,<br>0.384] | 0.003<br>[0.001,<br>0.004] | **0.001** | 0.727 | 1.259 | 36.868 | 0.114 |
| Fragmentation | 0.377<br>[0.347,<br>0.408] | 0.001 [-<br>0.000,<br>0.001] | 0.089 | 0.574 | 0.255 | 7.072 | 0.447<br>[0.407,<br>0.487] | 0.003<br>[0.002,<br>0.004] | **<0.001** | 0.619 | 1.043 | 35.204 | **0.001** |
|  | **Non-progressors, baseline Q8 = 2 (n=49)** |  |  |  |  |  | **Progressors, baseline Q8 = 2(n=19)** |  |  |  |  |  |  |
| Steps | 1555<br>[1293,<br>1817] | -11.12 [-<br>17.02,-<br>5.22] | **<0.001** | 0.765 | -0.822 | -37.18 | 1432<br>[959,<br>1905] | -28.67 [-<br>38.46,-<br>18.89] | **<0.001** | 0.827 | -1.583 | -104.104 | **0.001** |
| Cadence | 56.79<br>[52.05,<br>61.53] | -0.190 [-<br>0.288,-<br>0.093] | **<0.001** | 0.585 | -0.751 | -17.42 | 53.81<br>[43.54,<br>64.09] | -0.725 [-<br>0.958,-<br>0.493] | **<0.001** | 0.638 | -1.506 | -70.096 | **<0.001** |
| Intensity | 0.752<br>[0.682,<br>0.822] | -0.004 [-<br>0.006,-<br>0.003] | **<0.001** | 0.872 | **-0.921** | -27.924 | 0.674<br>[0.546,<br>0.802] | -0.008 [-<br>0.011,-<br>0.005] | **<0.001** | 0.905 | **-1.332** | **-58.604** | 0.018 |
| Similarity | 0.533<br>[0.487,<br>0.578] | -0.001 [-<br>0.001,-<br>0.000] | **0.017** | 0.794 | **-0.560** | -4.992 | 0.549<br>[0.447,<br>0.652] | -0.006 [-<br>0.009,-<br>0.004] | **<0.001** | 0.807 | **-1.193** | **-56.94** | **<0.001** |
| Variability | 0.477<br>[0.438,<br>0.516] | 0.000<br>[0.000,<br>0.001] | **0.001** | 0.857 | **0.548** | 4.524 | 0.480<br>[0.413,<br>0.548] | 0.003<br>[0.001,<br>0.005] | **<0.001** | 0.727 | **1.163** | **35.464** | **0.001** |
| Fragmentation | 0.571<br>[0.535,<br>0.607] | 0.002<br>[0.001,<br>0.004] | **0.001** | 0.664 | **0.987** | 20.956 | 0.569<br>[0.489,<br>0.648] | 0.006<br>[0.004,<br>0.009] | **<0.001** | 0.466 | **1.632** | **55.796** | **<0.001** |

**Figure 5.**
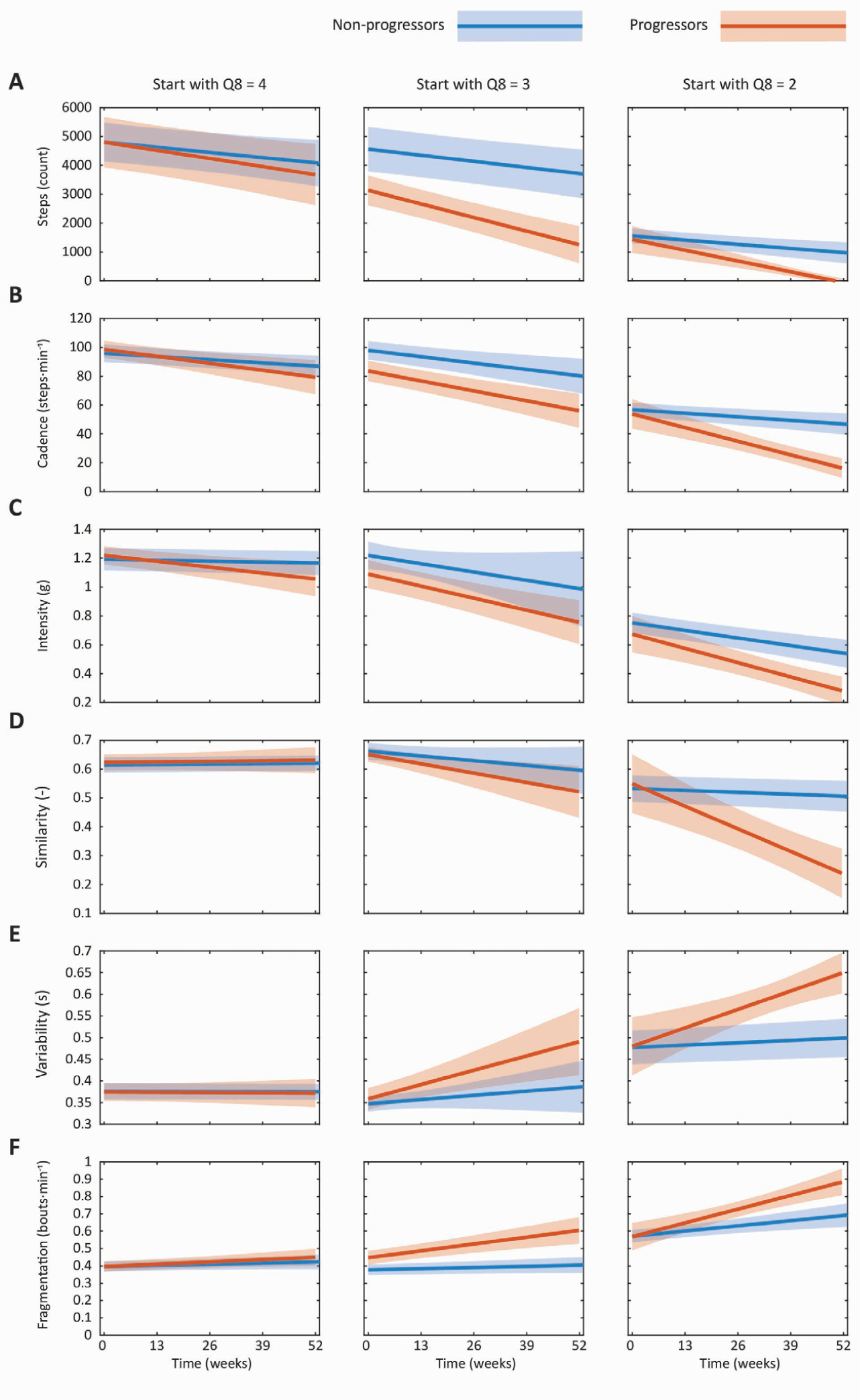
Population-level slopes of investigated gait metrics estimated using LMMs. Solid lines represent population-level mean trajectories of a gait metric (step counts (**A**), peak cadence (**B**), stride intensity (**C)**, stride pattern similarity (**D)**, stride duration variability (**E)**, and walking fragmentation (**F**)), while shaded areas represent their 95% confidence intervals. Trajectories depicted in blue represent participants that did not report gait decline on the ALSFRS-RSE over the observation period (non-progressors), while those depicted in red represent those who reported gait decline by at least one point on Q8 (progressors). Each gait metric was investigated separately in participants whose gait at enrollment was normal (left panels), experiencing early ambulation difficulties (middle panels), or required use of assistive devices (right panels).

As an example, we examined step counts. For non-progressors who started at Q8 = 4 and remained at 4 throughout the study, participants took an estimated 4805 steps/day at baseline (95% CI [4128, 5482]), which declined by 13.80 steps per week ([-25.14, -2.45]). Those who started with Q8 = 4 and declined over the study had a similar baseline of (4800 [3924, 5675]) but trended toward showing a steeper decline (-21.60 [-36.60, -6.60]).

For participants with early ambulation difficulties (Q8 = 3) who remained at 3 without decline, we observed significant decline only in step counts and peak cadence (both *p*=0.001), while those who started at Q8=3 and declined showed significant changes across all gait metrics (stride pattern similarity *p*=0.004, stride duration variability *p*=0.001, all other *p*<0.001). The rates of change between these two groups were significantly different for step counts (*p*<0.001) and walking fragmentation (*p=*0.001), suggesting that for those with early ambulation difficulty, gait deterioration is characterized by reduced steps and increased fragmentation (shorter bouts).

Among participants with baseline Q8 = 3 and no decline, the estimated baseline number of steps/day was 4560 ([3781, 5339]), with a decline of 16.25 steps per week (95%CI [-25.45, -7.06]). Those who started at Q8 = 3 and declined to <3 had a lower estimated baseline (3134 [2615, 3653]) and a steeper decline (36.23 [-47.30, -25.15]). The rates of change were significantly different between the those who remained a 3 and those who progressed to <3.

Finally, for participants walking with assistance (Q = 2) at baseline who did not decline, we observed significant change in each of the digital gait metrics, except stride pattern similarity (*p=*0.017). Those who started at Q = 2 and declined demonstrated significant change across each of the metrics, including similarity (*p<*0.001). Rates of decline in the two groups were significantly different across all metrics, except intensity (*p*=0.018), suggesting that participants walking with assistance (Q = 2) at baseline who progressed to a Q8 score of 0 or 1 (loss of ambulation) developed even slower steps, lower stride pattern similarity, greater stride duration variability, and more fragmented walking (shorter bouts) over the period of observation.

Among participants with baseline Q8 = 2 and no decline, the estimated baseline number of steps/day was 1555 (95% CI[1293, 1817]), with a decline of 11.12 steps per week ([-17.02, -5.22]). Those who started with Q8 = 2 and declined to <2 had a similar estimated baseline (1432 [959, 1905]), but a substantially steeper decline (-28.67 steps/week [-38.46, -18.89]). The rate of decline was significantly more rapid in people who became non-ambulatory (Q8 = 0 or 1; *p=*0.001).

### 3.4. Reliability and minimal detectable change

Test–retest reliability was calculated using two consecutive recording periods during the first year of data collection in which participants had a valid week of accelerometer data and reported no change in walking ability on Q8. Among individuals reporting normal gait (ALSFRS-RSE Q8 = 4), reliability ranged from moderate for peak cadence, intensity, similarity, and variability to good for step counts, while walking fragmentation demonstrated poor reliability. In participants reporting early ambulatory difficulties (Q8 = 3), most metrics demonstrated moderate reliability, with intensity showing good reliability (ICC = 0.780). The highest reliability estimates were observed among individuals requiring assistance with walking (Q8 = 2), where peak cadence demonstrated moderate reliability and all remaining gait metrics demonstrated good reliability, with intensity again demonstrating the highest reliability (ICC = 0.877) (**Table 5**).

**Table 5.** Reliability and minimal detectable change (%MDC95) for each of the digital gait metrics by Q8 score, n = number of observation pairs.

| Metric | ICC (95% CI) |  |  | %MDC95 |  |  |
| --- | --- | --- | --- | --- | --- | --- |
|  | 4<br>(n=197) | 3<br>(n=116) | 2<br>(n=246) | 4<br>(n=197) | 3<br>(n=116) | 2<br>(n=246) |
| Steps | 0.768<br>(0.704,<br>0.819) | 0.713<br>(0.596,<br>0.784) | 0.788<br>(0.741,<br>0.835) | 3767 | 2406 | 1337 |
| Cadence | 0.582<br>(0.480,<br>0.666) | 0.697<br>(0.582,<br>0.775) | 0.666<br>(0.592,<br>0.732) | 40.50 | 28.68 | 35.07 |
| Intensity | 0.619<br>(0.524,<br>0.698) | 0.780<br>(0.695,<br>0.841) | 0.877<br>(0.848,<br>0.905) | 0.443 | 0.294 | 0.293 |
| Similarity | 0.576<br>(0.472,<br>0.660) | 0.607<br>(0.471,<br>0.706) | 0.837<br>(0.797,<br>0.872) | 0.189 | 0.123 | 0.205 |
| Variability | 0.648<br>(0.559,<br>0.722) | 0.688<br>(0.571,<br>0.768) | 0.868<br>(0.835,<br>0.897) | 0.112 | 0.083 | 0.155 |
| Fragmentation | 0.497<br>(0.383,<br>0.594) | 0.679<br>(0.568,<br>0.767) | 0.751<br>(0.695,<br>0.804) | 0.226 | 0.117 | 0.230 |

MDC was estimated using SEM and ICC(2,1).^24^ For step counts, there was a decrease in the MDC value based on walking ability, indicating that those with lower scores on Q8 have less variability in their step counts. For peak cadence, stride intensity, stride similarity, stride duration variability, and walking fragmentation we again see a decrease in the MDC for those with early ambulation difficulties (Q8 = 3) compared to those reporting normal ambulation (Q8 = 4), indicating less variability in these metrics for individuals with the lower walking ability. However, findings for those requiring assistance (Q8 = 2) were heterogeneous. They demonstrated similar variability in intensity to those with early ambulatory difficulties, but greater variability in peak cadence compared to this group, and greater variability than both the normal gait and early ambulation difficulty groups in similarity, variability, and fragmentation (**Table 5**).

## 4. Discussion

The primary objective of this study was to develop and validate digital gait metrics obtained from an ankle-worn accelerometer to better understand changes in quantity and quality of gait in individuals with ALS. This work advances the development of fit-for-purpose digital gait endpoints that may complement traditional clinical assessments in both observational studies and interventional trials. Using a common analytical framework, we evaluated these endpoints comparing their cross-sectional associations with functional status, quantifying their rates of change over time, and estimating reliability and minimal detectable change. Building on our previous work,^13^ we analyzed several gait metrics, including step counts, peak cadence, stride intensity, stride pattern similarity, stride duration variability, and walking fragmentation, and compared these metrics to functional status captured by the ALSFRS-RSE. Our study helps address an important gap in the development of digital outcome measures in ALS. While prior work has shown that wearable sensors can accurately estimate gait characteristics,^13,27,28^ fewer studies have evaluated a broad set of metrics in free-living settings or examined how these metrics behave longitudinally across different ambulatory levels.

Importantly, our work supports convergent validity for accelerometry-based gait features, as all metrics were significantly associated with ALSFRS-RSE scores, with the strongest associations observed for the gross motor subdomain. These findings are consistent with prior studies demonstrating correlations between digital metrics and the ALSFRS-R.^29^ Importantly, several digital gait metrics were also highly intercorrelated. This suggests that although gait can be quantified by a complex interplay of spatiotemporal, kinematic, and kinetic parameters rather than just step counts or walking speed, these parameters capture related aspects of gait and may be meaningfully integrated into a composite score, or represented by a parsimonious, complementary subset of metrics.

The longitudinal analysis determined that participants showing declines in total step counts, peak cadence, stride intensity, and stride pattern similarity also experienced increased stride duration variability, walking fragmentation, and decreased functional scores. Many participants demonstrated greater relative change in the digital gait metrics as compared to total ALSFRS-RSE. These estimates were intended to provide a preliminary descriptive assessment of between-participant patterns but did not account for variability and therefore should not be interpreted as precise comparisons. Despite this, the observed patterns suggest that certain digital gait metrics may capture functional change more sensitively than the more global clinical rating scale in some individuals.

Perhaps not surprisingly, the analysis stratified by baseline walking impairment revealed that individuals with normal gait at baseline generally show stable performance over time, whereas those with early ambulation difficulties or a need for assistance demonstrated greater decline in performance across the digital gait metrics. In other words, if gait is unaffected at baseline, it is likely to remain normal over the observation period, while if gait has already begun to decline at baseline, the decline will continue. Similar patterns have been noted for bulbar function using quantitative motor speech analysis.^30^

At the same time, we found that several of the digital gait metrics appeared more sensitive to subtle gait changes, showing declines even when ALSFRS-RSE scores remained stable. Similarly to what has previously been observed in quantitative motor speech analyses, certain metrics may be more sensitive to change at different stages of ALS. Test–retest reliability ranged from moderate to good and revealed heterogeneous variability across gait metrics by level of gait impairment. Among individuals reporting normal gait, step counts demonstrated the highest reliability, whereas stride intensity showed the greatest reliability in those with early ambulatory difficulties or those requiring walking assistance. MDC values also varied by severity of gait impairment, with some gait metrics becoming more stable as gait impairment emerges, and others seeing more variability with advancing impairment, likely reflecting increased heterogeneity in compensatory strategies, assistive device use, and fluctuating mobility performance. Together, these findings support the clinical relevance of investigated gait metrics and further demonstrate that gait impairment in ALS is multidimensional and involves changes in walking volume, intensity, rhythm, consistency, and variability.^27,28^

The gait degradation represented by slowed peak cadence; decreased step counts, stride intensity, and stride similarity; and increased stride duration variability and fragmentation are consistent with prior reports of gait impairment in ALS as well as in other neurodegenerative disorders.^31,32^ For example, Dubbioso et al.^33^ reported increased variability in cadence and swing time in individuals with ALS, while Radovanović et al.^27^ observed longer gait cycle times and reduced stride lengths in individuals with both spinal- and bulbar-onset ALS. Our findings extend this work by demonstrating how multiple dimensions of gait change over time across individuals with varying levels of walking ability.^28^ We observed a decline in cadence, a metric of gait speed, over time and in correlation with decline in ALSFRS-RSE, confirming findings in prior studies.^28^ We found good correlations of ALSFRS-RSE with step counts and cadence, but lower correlations with stride pattern similarity or stride duration variability. Interestingly, a study in individuals with Huntington’s disease found stronger correlations between gait consistency measures and clinical outcomes than with step counts and cadence. While the reason for this difference is unknown, he driver of motor decline in Huntington’s is incoordination, which may cause primary failure in different gait subsystems.^31^ This highlights the importance of disease-specific clinical validation for digitally-derived mobility outcomes.

Finally, we found that increased walking fragmentation was also associated with ALS progression. Walking bout duration decreased over time, even in participants with stable total daily walking time, suggesting reduced capacity to sustain continuous walking as the disease progresses. Again, this further underscores the importance of deriving multiple complementary gait metrics from raw data to identify the key, most informative metrics of change in gait across different neurological diseases and across different stages of the same disease.

We acknowledge several limitations in our study. First, we used a conservative wear-time criterion of 16 hours for a valid day to improve data completeness, but this may have excluded individuals with lower adherence, lower functional abilities, discomfort with the device, or more variability in their daily routines.^34^ As a result, the analytic sample may overrepresent participants who were more engaged, with higher functional status, or more physically able, potentially limiting generalizability. Second, free-living data collection does not allow for a ground truth assessment of gait. Further analytical validation may improve the accuracy and interpretability of these digital gait metrics.^13^ Third, this dataset does not include gyroscopic data, thus cannot provide true gait velocity,^35,36^ which could be incorporated to future wDHT studies.^32^ Fourth, when calculating MDC, we relied on Q8 of the ALSFRS-RSE as the gold standard for decline in ambulatory status, however, it may be insensitive to small changes.^37^ This limitation could lead to a lower ICC, suggesting greater variability in the data than truly exists, leading to an overestimation of the MDC. Even so, the MDC estimation is useful, and we demonstrated that the MDC varies depending on severity of gait impairment – there does not appear to be a single MDC for all levels of impairment. While perhaps unexpected, this is consistent with previous findings of MDC for quantitative motor speech outcomes.^37,38^ Future studies of MDC could use shorter intervals for reassessment and employ alternative anchor questions.^39^ Finally, we did not directly assess the extent to which respiratory, cognitive or executive dysfunction could influence these gait metrics.^33^ Therefore, some of the variability in our digital metrics may reflect contributions from these non-motor factors and warrant further exploration in future studies.

## Conclusion

The development of objective, reliable, and sensitive digital gait metrics represents an important advancement in the measurement of gait in ALS. These findings underscore the potential of digital gait metrics to act as sensitive indicators of functional changes in plwALS and potentially endpoints in ALS trials. By complementing traditional clinical measures, these metrics may enable more precise tracking of disease progression and improve the evaluation of therapeutic interventions in both clinical care and clinical trials.

## 5. Declaration section

### 5.1. Ethics approval and consent to participate

The study was conducted in accordance with the ethical principles posited in the Declaration of Helsinki – Ethical Principles for Medical Research Involving Human Subjects. The Advarra Center for Institutional Review Board Intelligence gave ethical approval for this work (Pro00035442). Participants underwent the consent process and provided documentation of informed consent prior to any study procedures. There was no participant compensation.

### 5.2. Consent for publication

Not applicable.

### 5.3. Availability of data and materials

Data may be shared upon request and after review and approval by the owners of the data. Data may be requested through the ALS TDI ARC Data Commons (https://www.als.net/arc/data-commons/).

## 5.6. Acknowledgements

The authors acknowledge and thank Lori Chibnik, PhD for providing statistical guidance and contributing to discussions surrounding data interpretation.

## 5.7. Authors contributions

Concept and design – KB, NC, SM, KH, JB, MS

Data collection – AP, FGV

Statistical analysis – MS

Interpretation of results – KB, NC, SM, KH, JB, MS

Manuscript preparation – KB, MS

Preparation of tables and figures - MS

Critical review of manuscript – KB, NC, SM, KH, AP, FGV, JB, MS

Study supervision – FGV, JB

## 5.8. Disclosure of Delegation to Generative AI

The authors declare the use of generative AI in the research and writing process. According to the GAIDeT taxonomy (2025),^40^ the following tasks were delegated to GAI tools under full human supervision:

- Proofreading and editing

The GAI tool used was: Mass General Brigham AI Zone (aizone.mgb.org)

Responsibility for the final manuscript lies entirely with the authors.

GAI tools are not listed as authors and do not bear responsibility for the final outcomes.

Declaration submitted by: Katherine M. Burke

## References

1. Majmudar S, Wu J, Paganoni S. Rehabilitation in amyotrophic lateral sclerosis: why it matters. Muscle Nerve. 2014;50(1):4–13. doi:10.1002/mus.24202

2. Bello-Haas VD. Physical therapy for individuals with amyotrophic lateral sclerosis: current insights. Degener Neurol Neuromuscul Dis. 2018;8:45–54. doi:10.2147/DNND.S146949

3. Gil J, Funalot B, Verschueren A, et al. Causes of death amongst French patients with amyotrophic lateral sclerosis: a prospective study. Eur J Neurol. 2008;15(11):1245–1251. doi:10.1111/j.1468-1331.2008.02307.x

4. Garcia-Gancedo L, Kelly ML, Lavrov A, et al. Objectively Monitoring Amyotrophic Lateral Sclerosis Patient Symptoms During Clinical Trials With Sensors: Observational Study. JMIR MHealth UHealth. 2019;7(12):e13433. doi:10.2196/13433

5. van Unnik JWJ, Ing L, Oliveira Santos M, McDermott CJ, de Carvalho M, van Eijk RPA. Remote Monitoring of Amyotrophic Lateral Sclerosis Using Digital Health Technologies. Neurology. 2025;105(1):e213738. doi:10.1212/WNL.0000000000213738

6. Vieira FG, Venugopalan S, Premasiri AS, et al. A machine-learning based objective measure for ALS disease severity. NPJ Digit Med. 2022;5(1):45. doi:10.1038/s41746-022-00588-8

7. Johnson SA, Karas M, Burke KM, et al. Wearable device and smartphone data quantify ALS progression and may provide novel outcome measures. Npj Digit Med. 2023;6(1):1–10. doi:10.1038/s41746-023-00778-y

8. Cedarbaum JM, Stambler N, Malta E, et al. The ALSFRS-R: a revised ALS functional rating scale that incorporates assessments of respiratory function. BDNF ALS Study Group (Phase III). J Neurol Sci. 1999;169(1-2):13–21. doi:10.1016/s0022-510x(99)00210-5

9. Sukockienė E, Ferfoglia RI, Poncet A, Janssens JP, Allali G. Longitudinal Timed Up and Go Assessment in Amyotrophic Lateral Sclerosis: A Pilot Study. Eur Neurol. 2021;84(5):375–379. doi:10.1159/000516772

10. Two- and 6-minute walk tests assess walking capability equally in neuromuscular diseases. Neurology. Accessed July 29, 2024. https://www.neurology.org/doi/10.1212/WNL.0000000000002332

11. Inam S, Vucic S, Brodaty NE, Zoing MC, Kiernan MC. The 10-metre gait speed as a functional biomarker in amyotrophic lateral sclerosis. Amyotroph Lateral Scler Off Publ World Fed Neurol Res Group Mot Neuron Dis. 2010;11(6):558–561. doi:10.3109/17482961003792958

12. Musson LS, Mitic N, Leigh-Valero V, et al. The Use of Digital Devices to Monitor Physical Behavior in Motor Neuron Disease: Systematic Review. J Med Internet Res. 2025;27:e68479. doi:10.2196/68479

13. Straczkiewicz M, Burke KM, Carney KT, et al. Analytical and clinical validation of step counting method in people living with amyotrophic lateral sclerosis. Sci Rep. 2025;15(1):40769. doi:10.1038/s41598-025-24664-7

14. Micó-Amigo ME, Bonci T, Paraschiv-Ionescu A, et al. Assessing real-world gait with digital technology? Validation, insights and recommendations from the Mobilise-D consortium. J Neuroengineering Rehabil. 2023;20(1):78. doi:10.1186/s12984-023-01198-5

15. ALS Therapy Development Institute (ALS TDI). LS Research Collaborative (ARC) [Data set]. Published online 2023. 10.71944/C3NA-9124

16. Gupta AS, Patel S, Premasiri A, Vieira F. At-home wearables and machine learning sensitively capture disease progression in amyotrophic lateral sclerosis. Nat Commun. 2023;14(1):5080. doi:10.1038/s41467-023-40917-3

17. Schrack JA, Kuo PL, Wanigatunga AA, et al. Active-to-Sedentary Behavior Transitions, Fatigability, and Physical Functioning in Older Adults. J Gerontol A Biol Sci Med Sci. 2019;74(4):560–567. doi:10.1093/gerona/gly243

18. Choi L, Liu Z, Matthews CE, Buchowski MS. Validation of accelerometer wear and nonwear time classification algorithm. Med Sci Sports Exerc. 2011;43(2):357–364. doi:10.1249/MSS.0b013e3181ed61a3

19. Straczkiewicz M, Burke KM, Calcagno N, et al. Free-living monitoring of ALS progression in upper limbs using wearable accelerometers. J Neuroengineering Rehabil. 2024;21(1):223. doi:10.1186/s12984-024-01514-7

20. Koemel NA, Biswas RK, Ahmadi MN, et al. Minimum combined sleep, physical activity, and nutrition variations associated with lifeSPAN and healthSPAN improvements: a population cohort study. eClinicalMedicine. 2026;92:103741. doi:10.1016/j.eclinm.2025.103741

21. Shim J, Fleisch E, Barata F. Wearable-based accelerometer activity profile as digital biomarker of inflammation, biological age, and mortality using hierarchical clustering analysis in NHANES 2011–2014. Sci Rep. 2023;13(1):9326. doi:10.1038/s41598-023-36062-y

22. Ellis R, Nowell WB, Patel N, et al. Driving novel endpoints and study designs in amyotrophic lateral sclerosis: closer examination of the ALSFRS-R subdomains and a new definition of fast and slow progressors. medRxiv. Preprint posted online September 10, 2025:2025.08.21.25334108. doi:10.1101/2025.08.21.25334108

23. Schober P, Boer C, Schwarte LA. Correlation Coefficients: Appropriate Use and Interpretation. Anesth Analg. 2018;126(5):1763–1768. doi:10.1213/ANE.0000000000002864

24. Gates KE, Mefferd AS, Stipancic KL. Exploring Methodological Decisions for Calculating the Minimally Detectable Change in Dysarthria: Reliability, Statistics, and Standard Error of Measurement. J Speech Lang Hear Res JSLHR. 2025;68(8):3771–3788. doi:10.1044/2025_JSLHR-24-00899

25. Koo TK, Li MY. A Guideline of Selecting and Reporting Intraclass Correlation Coefficients for Reliability Research. J Chiropr Med. 2016;15(2):155–163. doi:10.1016/j.jcm.2016.02.012

26. Sedgwick P. Multiple hypothesis testing and Bonferroni’s correction. BMJ. 2014;349. doi:10.1136/bmj.g6284

27. Radovanović S, Milićev M, Perić S, Basta I, Kostić V, Stević Z. Gait in amyotrophic lateral sclerosis: Is gait pattern differently affected in spinal and bulbar onset of the disease during dual task walking? Amyotroph Lateral Scler Front Degener. 2014;15(7-8):488–493. doi:10.3109/21678421.2014.918148

28. Geronimo A, Martin AE, Simmons Z. Inertial sensing of step kinematics in ambulatory patients with ALS and related motor neuron diseases. J Med Eng Technol. 2021;45(6):486–493. doi:10.1080/03091902.2021.1922526

29. Holdom CJ, Pilkar R, Guo CC, et al. Identification of passive wrist-worn accelerometry outcomes for improved disease monitoring and trial design in motor neuron disease. eBioMedicine. 2025;117:105779. doi:10.1016/j.ebiom.2025.105779

30. Bingham IN, Norel R, Roitberg EG, et al. Listener effort measures clinically meaningful change of dysarthria in amyotrophic lateral sclerosis. Brain Commun. 2025;7(4):fcaf232. doi:10.1093/braincomms/fcaf232

31. Keren K, Busse M, Fritz NE, et al. Quantification of Daily-Living Gait Quantity and Quality Using a Wrist-Worn Accelerometer in Huntington’s Disease. Front Neurol. 2021;12:719442. doi:10.3389/fneur.2021.719442

32. Zhang W, Ling Y, Chen Z, et al. Wearable sensor-based quantitative gait analysis in Parkinson’s disease patients with different motor subtypes. Npj Digit Med. 2024;7(1):169. doi:10.1038/s41746-024-01163-z

33. Dubbioso R, Spisto M, Hausdorff JM, et al. Cognitive impairment is associated with gait variability and fall risk in amyotrophic lateral sclerosis. Eur J Neurol. 2023;30(10):3056–3067. doi:10.1111/ene.15936

34. Straczkiewicz M, Burke KM, Calcagno N, et al. Short prescribed exercises can quantify upper limb functioning in neurodegenerative disease. J Neuroengineering Rehabil. 2026;23(1):28. doi:10.1186/s12984-025-01829-z

35. Rabbia M, Guridi Ormazabal M, Staunton H, et al. Stride Velocity 95th Centile Detects Decline in Ambulatory Function Over Shorter Intervals than the 6-Minute Walk Test or North Star Ambulatory Assessment in Duchenne Muscular Dystrophy. J Neuromuscul Dis. 2024;11(3):701–714. doi:10.3233/JND-230188

36. Poleur M, Willekens B, Degos B, et al. Stride-level measurement of gait as an early sensitive marker of disability progression in ambulatory patients with multiple sclerosis. EClinicalMedicine. 2026;93:103823. doi:10.1016/j.eclinm.2026.103823

37. Stipancic KL, Yunusova Y, Berry JD, Green JR. Minimally Detectable Change and Minimal Clinically Important Difference of a Decline in Sentence Intelligibility and Speaking Rate for Individuals With Amyotrophic Lateral Sclerosis. J Speech Lang Hear Res. 2018;61(11):2757–2771. doi:10.1044/2018_JSLHR-S-17-0366

38. Stipancic KL, Tjaden K. Minimally Detectable Change of Speech Intelligibility in Speakers With Multiple Sclerosis and Parkinson’s Disease. J Speech Lang Hear Res JSLHR. 2022;65(5):1858–1866. doi:10.1044/2022_JSLHR-21-00648

39. Kamper SJ, Maher CG, Mackay G. Global Rating of Change Scales: A Review of Strengths and Weaknesses and Considerations for Design. J Man Manip Ther. 2009;17(3):163. doi:10.1179/jmt.2009.17.3.163

40. GAIDeT (Generative AI Delegation Taxonomy): A taxonomy for humans to delegate tasks to generative artificial intelligence in scientific research and publishing: Accountability in Research: Vol 0, No 0 - Get Access. Accessed January 1, 2026. https://www.tandfonline.com/doi/full/10.1080/08989621.2025.2544331

