## Supplementary Materials for "Quantifying longitudinal gait changes in ALS using wearable digital health technology metrics"

### 1 26-week time window

#### 1.1 Population-level trajectories

**Table S1.** Model coefficients for digital gait metrics and functional scores, represented by the total and gross motor sub-domain scores on the ALSFRS-RSE. Model slopes express weekly change in digital metrics and survey scores. Standardized and relative changes are expressed in SD and percentage change per 26 weeks, respectively.

| Outcome | Intercept | Slope | *p*-value | R-squared | Standardized change (SD) | Relative change (%) |
|---|---|---|---|---|---|---|
| Steps | 3333 [3007, 3658] | -29.02 [-42.02, -16.02] | <0.001 | 0.913 | -0.589 | -22.646 |
| Cadence | 80.88 [76.81, 84.96] | -0.485 [-0.681, -0.288] | <0.001 | 0.817 | -0.871 | -15.574 |
| Intensity | 1.024 [0.9743, 1.074] | -0.005 [-0.007, -0.003] | <0.001 | 0.845 | -0.771 | -13.676 |
| Similarity | 0.600 [0.58, 0.620] | -0.002 [-0.003, -0.000] | 0.011 | 0.834 | -0.546 | -6.838 |
| Variability | 0.404 [0.387, 0.421] | 0.001 [0.001, 0.002] | 0.002 | 0.890 | 0.599 | 8.944 |
| Fragmentation | 0.465 [0.443, 0.487] | 0.003 [0.001, 0.004] | <0.001 | 0.762 | 0.827 | 14.586 |
| Q7—9 | 9.23 [8.90, 9.56] | -0.037 [-0.047, -0.027] | <0.001 | 0.976 | -0.857 | -10.452 |
| Q1—12 | 40.54 [39.79, 41.29] | -0.108 [-0.131, -0.084] | <0.001 | 0.978 | -1.062 | -6.89 |

#### 1.2 Participant-level comparison

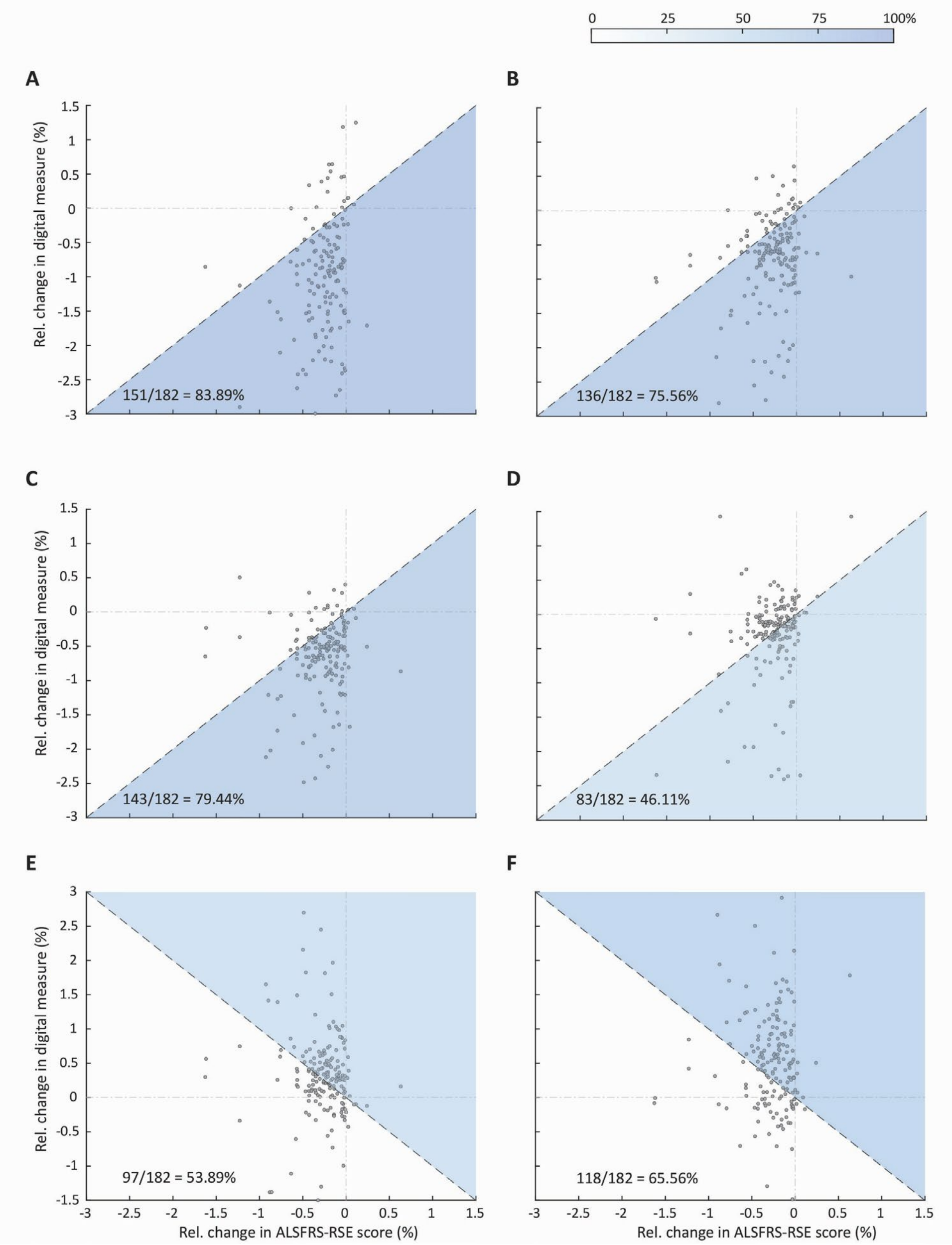


**Figure S1. Point estimates for participant-specific relative change per week in digital gait metrics compared to corresponding relative change in ALSFRS-RSE total score within a 26-week window.** Each panel corresponds to one metric: step counts (**A**), peak cadence (**B**), stride intensity (**C**), stride pattern similarity (**D**), stride duration variability (**E**), and walking fragmentation (**F**). Each point represents the relative change in the digital metric and ALSFRS-RSE total score in one participant; dashed lines correspond to identical relative change observed in the digital gait metric and the ALSFRS-RSE total score. The shaded sections highlight individuals with relative change in the digital metric exceeding the relative change in their ALSFRS-RSE total score. Tones of blue indicate the percentage of participants with greater relative change exhibited by a digital metric, with darker blues indicating higher percentage.

#### 1.3 Stratification by self-reported disease status and its change

**Table S2.** Comparison between longitudinal changes in gait metrics between individuals reporting no decline (non-progressors) and those reporting decline by at least one point (progressors) on ALSFRS-RSE Q8. Model slopes express weekly change in digital metrics and survey scores. Standardized and relative changes are expressed in SD and percentage change per 26 weeks, respectively.

| Metric | Intercept | Slope | Slope *p*-value | R-squared | Standardized change (SD) | Relative change (%) | Intercept | Slope | Slope *p*-value | R-squared | Standardized change (SD) | Relative change (%) | Slope difference p-value |
|---|---|---|---|---|---|---|---|---|---|---|---|---|---|
| | | **Non-progressors, baseline Q8 = 4 (n=53)** | | | | | **Progressors, baseline Q8 = 4 (n=11)** | | | | | | |
| Steps | 4751 [3939, 5563] | -24.15 [-72.33, 24.02] | 0.323 | 0.795 | -0.414 | -13.208 | 4798 [3967, 5629] | -33.82 [-70.66, 3.02] | 0.072 | 0.869 | -0.587 | -18.33 | 0.624 |
| Cadence | 96.96 [88.13, 105.8] | -0.417 [-0.961, 0.126] | 0.131 | 0.610 | -0.735 | -11.18 | 100.2 [94.12, 106.3] | -0.645 [-1.06, -0.228] | **0.003** | 0.736 | -1.430 | -16.718 | 0.566 |
| Intensity | 1.19 [1.09, 1.29] | -0.000 [-0.006, 0.006] | 0.892 | 0.625 | 0.015 | -0.91 | 1.24 [1.17, 1.31] | -0.006 [-0.010, -0.003] | **0.001** | 0.309 | -1.358 | -13.494 | 0.103 |
| Similarity | 0.610 [0.568, 0.653] | -0.000 [-0.003, 0.003] | 0.937 | 0.652 | 0.033 | -0.546 | 0.632 [0.601, 0.663] | -0.001 [-0.003, 0.000] | 0.150 | 0.419 | -0.646 | -5.486 | 0.598 |
| Variability | 0.379 [0.351, 0.407] | -0.000 [-0.002, 0.002] | 0.910 | 0.681 | -0.181 | -0.676 | 0.368 [0.347, 0.390] | 0.001 [-0.000, 0.002] | 0.133 | 0.530 | 0.600 | 6.084 | 0.395 |
| Fragmentation | 0.405 [0.355, 0.456] | 0.001 [-0.003, 0.005] | 0.720 | 0.568 | 0.266 | 4.602 | 0.391 [0.360, 0.422] | 0.002 [-0.000, 0.005] | 0.090 | 0.375 | 0.911 | 14.04 | 0.485 |
| | | **Non-progressors, baseline Q8 = 3 (n=33)** | | | | | **Progressors, baseline Q8 = 3 (n=17)** | | | | | | |
| Steps | 4437 [3702, 5172] | -2.945 [-42.434, 36.544] | 0.882 | 0.779 | -0.075 | -1.716 | 3222 [2697, 3747] | -51.61 [-78.30, -24.93] | **<0.001** | 0.815 | -1.659 | -41.6 | 0.010 |
| Cadence | 97.41 [91.33, 103.5] | -0.407 [-1.012, 0.198] | 0.184 | 0.743 | -1.090 | -10.868 | 85.37 [77.36, 93.37] | -0.799 [-1.288, -0.309] | **0.002** | 0.810 | -1.831 | -24.336 | 0.131 |

| | | | | | | | | | | | | |
|---|---|---|---|---|---|---|---|---|---|---|---|---|
| Intensity | 1.19 [1.10, 1.28] | -0.001 [-0.008, 0.005] | 0.740 | 0.866 | -0.248 | -0.091 | 1.10 [0.993, 1.20] | -0.008 [-0.014, -0.002] | 0.010 | 0.906 | -1.263 | -18.798 | 0.118 |
| Similarity | 0.652 [0.6214, 0.6827] | 0.000 [-0.001, 0.002] | 0.540 | 0.701 | 0.300 | 0.071 | 0.650 [0.616, 0.684] | -0.003 [-0.005, 0.000] | 0.053 | 0.618 | -1.410 | -10.894 | 0.117 |
| Variability | 0.356 [0.336, 0.376] | -0.001 [-0.002, 0.000] | 0.101 | 0.713 | -0.874 | -0.229 | 0.360 [0.330, 0.389] | 0.003 [0.000, 0.005] | 0.038 | 0.866 | 1.468 | 19.292 | 0.062 |
| Fragmentation | 0.387 [0.352, 0.421] | -0.001 [-0.003, 0.000] | 0.150 | 0.520 | -0.806 | -0.322 | 0.443 [0.401, 0.485] | 0.004 [0.001, 0.007] | 0.008 | 0.820 | 1.679 | 24.752 | 0.030 |
| | | **Non-progressors, baseline Q8 = 2 (n=60)** | | | | | **Progressors, baseline Q8 = 2 (n=8)** | | | | | | |
| Steps | 1695 [1394, 1995] | -24.85 [-38.92, -10.77] | **0.001** | 0.752 | -1.130 | -38.116 | 1397 [945.1, 1849] | -28.65 [-39.97, -17.33] | **<0.001** | 0.882 | -1.483 | -53.3 | 0.754 |
| Cadence | 59.21 [53.78, 64.64] | -0.435 [-0.774, -0.095] | 0.012 | 0.643 | -0.964 | -19.084 | 50.07 [40.1, 60.05] | -0.361 [-0.772, 0.049] | 0.083 | 0.432 | -1.169 | -18.772 | 0.946 |
| Intensity | 0.776 [0.701, 0.851] | -0.007 [-0.010, -0.004] | **<0.001** | 0.910 | -1.329 | -23.504 | 0.647 [0.516, 0.777] | -0.006 [-0.009, -0.002] | **0.006** | 0.946 | -1.294 | -22.464 | 0.663 |
| Similarity | 0.551 [0.506, 0.596] | -0.003 [-0.005, -0.001] | 0.010 | 0.842 | -0.812 | -13.702 | 0.511 [0.417, 0.605] | -0.003 [-0.008, 0.002] | 0.267 | 0.946 | -0.812 | -15.028 | 0.907 |
| Variability | 0.4593 [0.4177, 0.501] | 0.002 [0.001, 0.004] | **0.006** | 0.903 | 0.724 | 13.104 | 0.494 [0.427, 0.560] | 0.003 [-0.001, 0.006] | 0.102 | 0.844 | 1.203 | 13.39 | 0.782 |
| Fragmentation | 0.551 [0.511, 0.591] | 0.005 [0.002, 0.008] | **0.002** | 0.845 | 1.381 | 21.892 | 0.599 [0.517, 0.680] | 0.003 [-0.002, 0.008] | 0.295 | 0.628 | 1.041 | 11.362 | 0.577 |

### 2 13-week time window

#### 2.1 Population-level trajectories

**Table S3.** Model coefficients for digital gait metrics and functional scores, represented by the total and gross motor sub-domain scores on the ALSFRS-RSE. Model slopes express weekly change in digital metrics and survey scores. Standardized and relative changes are expressed in SD and percentage change per 13 weeks, respectively.

| Outcome | Intercept | Slope | Slope *p*-value | R-squared | Standardized change (SD) | Relative change (%) |
|---|---|---|---|---|---|---|
| Steps | 3363 [3017, 3710] | -38.70 [-60.69, -16.70] | 0.001 | 0.954 | -0.984 | -14.963 |
| Cadence | 80.84 [76.55, 85.14] | -0.490 [-0.844, -0.136] | 0.007 | 0.946 | -1.065 | -7.878 |
| Intensity | 1.019 [0.969, 1.070] | -0.004 [-0.008, -0.001] | 0.014 | 0.973 | -0.748 | -5.603 |
| Similarity | 0.597 [0.576, 0.618] | -0.001 [-0.003, 0.001] | 0.386 | 0.936 | -0.303 | -1.82 |
| Variability | 0.405 [0.388, 0.422] | 0.001 [-0.000, 0.002] | 0.068 | 0.968 | 0.613 | 3.692 |
| Fragmentation | 0.469 [0.446, 0.492] | 0.002 [-0.001, 0.004] | 0.151 | 0.926 | 0.726 | 4.953 |
| Q7—9 | 9.233 [8.91, 9.56] | -0.037 [-0.052, -0.021] | <0.001 | 0.989 | -0.808 | -5.148 |
| Q1—12 | 40.56 [39.82, 41.31] | -0.114 [-0.146, -0.081] | <0.001 | 0.984 | -1.145 | -3.64 |

#### 2.2 Participant-level comparison

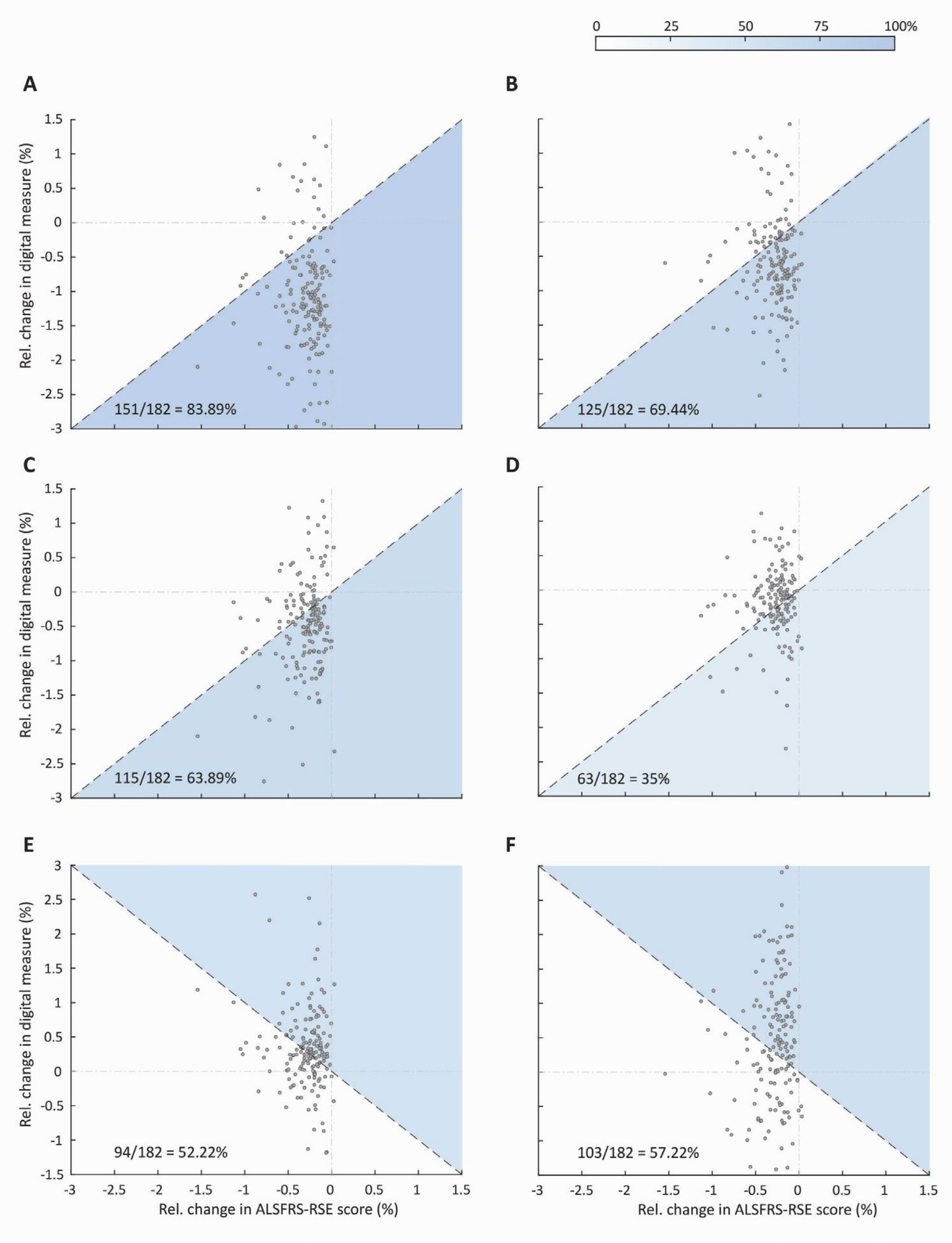


**Figure S2. Point estimates for participant-specific relative change per week in digital gait metrics compared to corresponding relative change in ALSFRS-RSE total score within a 13-week window.** Each panel corresponds to one metric: step counts (**A**), peak cadence (**B**), stride intensity (**C**), stride pattern similarity (**D**), stride duration variability (**E**), and walking fragmentation (**F**). Each point represents the relative change in the digital metric and ALSFRS-RSE total score in one participant; dashed lines correspond to identical relative change observed in the digital gait metric and the ALSFRS-RSE total score. The shaded sections highlight individuals with relative change in the digital metric exceeding the relative change in their ALSFRS-RSE total score. Tones of blue indicate the percentage of participants with greater relative change exhibited by a digital metric, with darker blues indicating higher percentage.

#### 2.3 Stratification by self-reported disease status and its change

**Table S4.** Comparison between longitudinal changes in gait metrics between individuals reporting no decline (non-progressors) and those reporting decline by at least one point (progressors) on ALSFRS-RSE Q8. Model slopes express weekly change in digital metrics and survey scores. Standardized and relative changes are expressed in SD and percentage change per 13 weeks, respectively.

| **Metric** | **Intercept** | **Slope** | **Slope *p*-value** | **R-squared** | **Standardized change (SD)** | **Relative change (%)** | **Intercept** | **Slope** | **Slope *p*-value** | **R-squared** | **Standardized change (SD)** | **Relative change (%)** | **Slope difference p-value** |
|---|---|---|---|---|---|---|---|---|---|---|---|---|---|
| | | **Non-progressors, baseline Q8 = 4 (n=59)** | | | | | **Progressors, baseline Q8 = 4 (n=5)** | | | | | | |
| Steps | 4882 [3985, 5780] | -77.66 [-167.65, 12.32] | 0.090 | 0.809 | -2.174 | -20.67 | 4842 [3936, 5748] | -34.66 [-82.83, 13.51] | 0.156 | 0.922 | -0.644 | -9.308 | 0.579 |
| Cadence | 98 [87.13, 108.9] | -0.881 [-2.16, 0.405] | 0.177 | 0.910 | -2.185 | -11.687 | 101.1 [94.67, 107.4] | -0.675 [-1.141, -0.209] | **0.005** | 0.903 | -1.610 | -8.684 | 0.902 |
| Intensity | 1.19 [1.07, 1.30] | -0.000 [-0.014, 0.014] | 0.965 | 0.839 | -0.550 | -0.325 | 1.23 [1.16, 1.30] | -0.004 [-0.011, 0.002] | 0.204 | 0.841 | -0.889 | -4.602 | 0.510 |
| Similarity | 0.607 [0.557, 0.657] | 0.000 [-0.005, 0.006] | 0.885 | 0.743 | -0.258 | 0.897 | 0.628 [0.594, 0.661] | 0.000 [-0.002, 0.002] | 0.793 | 0.818 | 0.210 | 0.598 | 0.913 |
| Variability | 0.379 [0.347, 0.412] | 0.000 [-0.003, 0.004] | 0.884 | 0.909 | 0.540 | 0.91 | 0.370 [0.347, 0.393] | 0.000 [-0.001, 0.002] | 0.772 | 0.911 | -0.033 | 0.754 | 0.938 |

| | | | | | | | | | | | | | |
|---|---|---|---|---|---|---|---|---|---|---|---|---|---|
| Fragmentation | 0.411 [0.348, 0.474] | 0.001 [-0.008, 0.009] | 0.896 | 0.922 | 0.892 | 1.677 | 0.396 [0.365, 0.427] | 0.000 [-0.002, 0.002] | 0.980 | 0.911 | 0.015 | 0.078 | 0.900 |
| | | **Non-progressors, baseline Q8 = 3 (n=36)** | | | | | **Progressors, baseline Q8 = 3 (n=14)** | | | | | | |
| Steps | 4520 [3704, 5336] | -32.50 [-105.80, 40.79] | 0.377 | 0.872 | -0.978 | -9.347 | 3253 [2679, 3827] | -69.70 [-108.59, -30.82] | **0.001** | 0.901 | -2.374 | -27.859 | 0.371 |
| Cadence | 97.08 [90.98, 103.2] | -0.583 [-1.76, 0.596] | 0.325 | 0.969 | -1.654 | -7.8 | 85.06 [77.12, 93.01] | -0.981 [-1.54, -0.420] | **0.001** | 0.861 | -2.349 | -14.989 | 0.229 |
| Intensity | 1.18 [1.09, 1.27] | -0.001 [-0.010, 0.008] | 0.896 | 0.948 | -0.143 | -0.637 | 1.08 [0.978, 1.19] | -0.009 [-0.014, -0.003] | **0.002** | 0.940 | -1.505 | -10.179 | 0.103 |
| Similarity | 0.650 [0.618, 0.682] | 0.001 [-0.002, 0.004] | 0.646 | 0.657 | 0.503 | 1.43 | 0.642 [0.6055, 0.678] | -0.004 [-0.010, 0.002] | 0.154 | 0.945 | -1.483 | -8.099 | 0.200 |
| Variability | 0.356 [0.335, 0.377] | -0.001 [-0.003, 0.001] | 0.457 | 0.511 | -0.824 | -2.691 | 0.365 [0.333, 0.398] | 0.003 [-0.000, 0.005] | 0.051 | 0.945 | 1.895 | 9.555 | 0.118 |
| Fragmentation | 0.386 [0.347, 0.426] | -0.001 [-0.006, 0.003] | 0.483 | 0.980 | -0.965 | -4.979 | 0.441 [0.398, 0.484] | 0.007 [0.003, 0.011] | 0.002 | 0.632 | 2.873 | 19.37 | 0.019 |
| | | **Non-progressors, baseline Q8 = 2 (n=63)** | | | | | **Progressors, baseline Q8 = 2 (n=5)** | | | | | | |

| | | | | | | | | | | | | | |
|---|---|---|---|---|---|---|---|---|---|---|---|---|---|
| Steps | 1671 [1370, 1973] | -12.80 [-39.81, 14.21] | 0.350 | 0.819 | -0.727 | -9.958 | 1401 [958.5, 1843] | -28.45 [-57.02, 0.12] | 0.051 | 0.974 | -1.212 | -26.403 | 0.499 |
| Cadence | 58.5 [52.95, 64.05] | -0.147 [-0.780, 0.486] | 0.646 | 0.851 | -0.262 | -3.263 | 49.15 [38.85, 59.45] | -0.176 [-1.023, 0.672] | 0.678 | 0.727 | -1.075 | -4.641 | 0.970 |
| Intensity | 0.783 [0.705, 0.860] | -0.007 [-0.012, -0.002] | **0.005** | 0.969 | -1.275 | -12.22 | 0.646 [0.518, 0.774] | -0.007 [-0.012, -0.001] | 0.016 | 0.974 | -1.689 | -13.871 | 0.918 |
| Similarity | 0.553 [0.506, 0.600] | -0.003 [-0.007, 0.001] | 0.174 | 0.944 | -0.677 | -6.825 | 0.507 [0.412, 0.603] | -0.001 [-0.008, 0.006] | 0.766 | 0.953 | 0.069 | -2.769 | 0.651 |
| Variability | 0.458 [0.416, 0.500] | 0.002 [-0.001, 0.004] | 0.211 | 0.935 | 0.532 | 4.368 | 0.489 [0.425, 0.553] | 0.003 [-0.002, 0.009] | 0.230 | 0.957 | 1.523 | 9.217 | 0.475 |
| Fragmentation | 0.559 [0.519, 0.598] | 0.001 [-0.002, 0.005] | 0.470 | 0.829 | 0.290 | 3.081 | 0.599 [0.510, 0.689] | 0.004 [-0.008, 0.015] | 0.526 | 0.782 | 0.537 | 7.943 | 0.708 |
